# Optimal control and cost-effectiveness analysis of a fractional order drug-resistant malaria transmission model with recovered carriers

**DOI:** 10.1101/2025.05.22.25328145

**Authors:** Amit Kumar Saha, Shikha Saha, Chandra Nath Podder

## Abstract

Malaria is still a life threatening parasitic disease due to the change in environmental and socio-economic conditions. This paper introduces a novel mathematical model to study the impact of drug-resistant strain, recovered-carrier, and relapse on malaria dynamics by implementing Caputo-Fabrizio fractional order derivative (CFFOD). We begin by presenting theoretical results that are derived for our model. We also derive the expression for the control reproduction number and investigate the equilibria of the proposed model. Theoretical analysis guarantees the presence of a unique solution to the suggested model. We use a combination of Runge-Kutta fourth order method and three-step Adams–Bashforth scheme to obtain numerical solution. Additionally, to control the spread of malaria and also to understand the epidemiological characteristics and public health indicators of malaria, three control variables are introduced in this model. Finally, a rigorous cost-effectiveness analysis is performed to determine the most economical control variable. The possibility of backward bifurcation is suggested by numerical simulation, which reveal the coexistence of a stable disease free equilibrium (DFE) and a stable endemic equilibrium point (EEP). According to the simulation, it is also observed that due to the existence of recovered carriers in the population the disease tends to linger. The study further reveals that an increase in the drug-resistant strain increase the disease-related complexity. A thorough sensitivity analysis is also carried out to identify the most sensitive parameters that control the basic reproduction number. According to our findings, vaccination control is the most cost-effective strategy to reduce disease burden.

## 1. Introduction

Malaria is an ancient parasitic disease that poses serious health, social, and economic burden. Humans are infected with malaria through the bite of a female Anopheles mosquito carrying the Plasmodium parasite. [1]. Out of the four species of plasmodium (plasmodium falciparum, plasmodium vivax, plasmodium malariae, and plasmodium ovale) that infect humans, plasmodium falciparum and plasmodium vivax are the deadliest [2]. Malaria is mainly found in tropical and sub-tropical regions in the world occurring mostly in sub-Saharan Africa and South Asia [3]. Its existence has long been known, but still, it remains a major public health concern around the world. Even though malaria is curable and preventable, it has been endemic in many African countries. It affects children under 5, non-immune adults, and pregnant women, especially in countries where it is endemic. Malaria outbreak and its spread are influenced by different factors such as temperature, rainfall, and humidity [3]. Malaria infected individuals experience chills, headaches, fever, and other flu-like symptoms [4]. Unfortunately, if left untreated, the disease may develop severe illness and the patient may die [5]. According to CDC, in 2020, approximately 241 million cases occurred along with an estimated 627,000 deaths [6].

To reduce the spread of malaria, preventive measures should be taken seriously. Malaria prevention measure includes insecticide-treated bed nets which proved to be very much effective. However, mosquitoes have managed to evolve to develop resistance against insecticide-treated bed nets. Other measures are insect-repellent creams and DDT spray [7]. The creams are also very effective but at the same time, they are very costly. The draining of standing water and other water lands is also an effective way to prevent malaria [8]. Malaria treatment includes artemisinin-based therapy. Individuals diagnosed with malaria are treated based on categories as having either complicated or uncomplicated malaria [9]. Oral anti-malarials are suggested for patients with mild malaria. However patients diagnosed with severe malaria should be treated with intravenous antimalarial therapy. To prevent malaria, antimalarial drugs can also be recommended. Malaria drugs include Chloroquine, Doxycycline, Mefloquine, Primaquine, and Tafenoquine [10]. Yet one of the greatest threats to controlling malaria is the emergence of drug-resistance strains, which increases malaria-related morbidity and mortality. However, only Plasmodium falciparum and P. vivax, of the four human malaria parasite species, have been found to have antimalarial drug resistance [11]. Hence, to advance the fight against malaria, better tools are still needed. This necessitates the development of vaccines which can be another significant preventive measure. Over the past few decades, a tremendous amount of effort has been put forth in the pursuit of effective vaccines. Finally, RTS,S is the first malaria vaccine approved by WHO [12]. After the artemisinin-based therapy, drugs, bed nets, and repellents (non-pharmaceutical interventions), RTS,S is the new best tool to fight against malaria. Mathematical models always represent simplified versions of reality, yet, the main features of the disease can still be captured. They can also provide important information about disease transmission and the complex mechanism of different control strategies to reduce the disease burden. Further, they can predict the disease outcome and hence can provide useful information about public health intervention policies. Similarly, to reduce the deadly impact of malaria, various mathematical models considering the epidemiological features of malaria have been constructed and rigorously studied. The first epidemiological model to describe malaria was developed by Ross in the form of a deterministic compartmental model [13]. According to Ross, malaria can be removed from the community if the number of mosquitoes can be brought under a certain quantity. Later on, by modifying the model including super-infection, Macdonald [14] showed that reduction in mosquito populations has little effect in areas where intense transmission occurs. In [15], the authors modified the model in [14] by adding various features of malaria such as an incubation period for mosquitoes and immunity for humans. The authors in [16], proposed a model where hosts follow the SEIRS pattern and vectors follow the SEI pattern. Furthermore, Chitnis et al [17] reformulated the model described in Ngwa and Shu, [16] by considering that recovered people may not be fully recovered. In [18], the authors explored how the development of insecticide resistance in the vector population influences the spread and persistence of malaria. They concluded that ITNs and IRS coverage are required to effectively control the disease. Esteva et al. [19] studied a malaria transmission model considering resistance strain. They discussed various strategies for effectively controlling drug resistance, such as, threshold reduction, optimal drug usage, and inception of combination therapies. There are many more mathematical models regarding malaria. However, all these mathematical models consider integer order derivatives and hence have limitations due to their local nature [20]. The classical integer order epidemic models can not explain the memory and the learning mechanism of the host or vector on disease transmission. But in epidemiology, it’s important to predict the future and to understand how to control its progress [21]. The fractional order model can give useful information about their previous states which will be helpful to incorporate memory and hence they can predict the future more convincingly [22]. Fractional order calculus is the generalization of the classical integer order calculus and the order of the fractional order operator is considered as the memory index [20]. Fractional order model has also the ability to address the real world phenomena more accurately. For this reason, fractional-order equations are frequently used even though integer-order equations are successful [23]. Another key benefit of fractional derivative over classical integer order derivative is that they can provide critical information about the dynamics of the system between two distinct points. Therefore fractional order derivative approaches in the model formulation are suggested. Authors from different fields have made significant contribution in mathematical modeling of infectious disease by incorporating fractional-order differential equations. The most common are the Caputo, the Riemann-Liovile and the Grunwald-Letnikoy formulations. The Caputo fractional derivative (CFD) was proposed by Caputo which takes into account the interactions within the past and the problems with non-local properties. But, sometimes the kernel defined in CFD becomes singular and hence CFD can not always describe the memory effect accurately. Recently, a new fractional order derivative, considering non-singular kernel, is presented by Caputo and Fabrizio. Many authors have applied this newly introduced Caputo-Fabrizio fractional order derivative (CFFOD) in modeling various real-world phenomena from different fields. Authors in [23], considered a model to understand the expansion and extension of computer viruses considering the Caputo-Fabrizio fractional differential equation (CFFDE). In [24], authors proved the existence and uniqueness of solutions for water flow dynamics within a closed reservoir by formulating a CFFOD model. Further, using CFFOD, authors in [20] evaluated the impact of vaccination, endogenous re-activation, and exogenous re-infection on TB dynamics. In their study [25], Gizaw et. al. introduced a novel fractional order model by incorporating variability in temperature and rainfall to determine the effect of temperature and rainfall on malaria transmission dynamics. In [26], the authors presented a novel fractional order Malaria model incorporating optimal control. Two control variables were considered for treatment to be successful. Pinto et. al [7] considered a fractional model to study malaria transmission dynamics and its health & socioeconomic burden.

Inspired by the effectiveness of the CF fractional-order derivative observed in prior research, we propose a novel mathematical model to study the dynamics of malaria transmission by incorporating CF fractional order derivatives. It is known from the literature that malaria is not always fully curable and there are some drug-resistance cases also. But none of the articles discussed above considered these two features simultaneously while we have included both of them in our model. We have also considered vaccination to understand the effect of vaccines on malaria control. We have further considered treatment for both drug resistance and drug-sensitive cases which is also a new feature of the model.

The entire paper is decorated as follows. In section 2, we have discussed some basic preliminaries regarding Caputo-Fabrizio fractional order derivative. Section 3 deals with the formulation of the model and the discussion about the model variables and parameters. Section 4 deals with the theoretical analysis of the model. In section 5, three-step Adams-Bashforth numerical scheme has been discussed. Numerical simulation is carried out in section 7. The main findings of this study have been discussed and conclusion has been drawn in section 8.

## 2. Basic preliminaries

**Definition 1**. *[27, 21] Let f* ∈ *H*^1^(*a, b*) *and ρ* ∈ (0, 1). *Then the CFFOD is defined as*

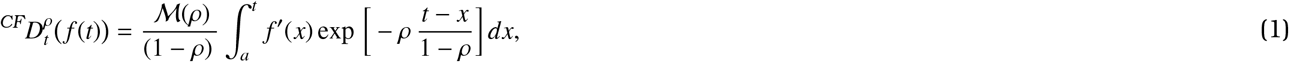

*where ℳ* (*ρ*) *has the properties that ℳ* (0) = *ℳ* (1) = 1.

**Definition 2**. *[27, 21] Let* 0 *< ρ <* 1. *The fractional integral of order ρ of a function f is defined by*

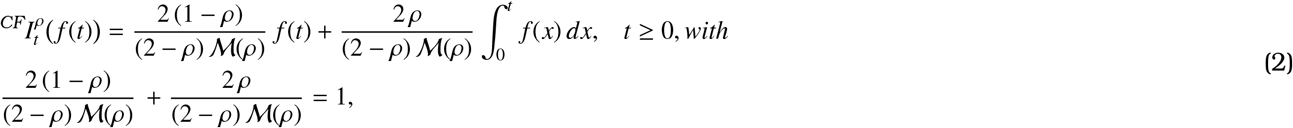

Using *ℳ(p)=2/2-p*, Losada and Nieto proposed the new Caputo derivative and its corresponding integral as follows:

**Definition 3**. *[27, 22] Let ρ* ∈ (0, 1). *Then the CFFD of order ρ is defined by*

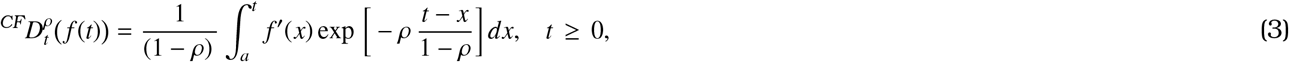

and its fractional integration is defined as

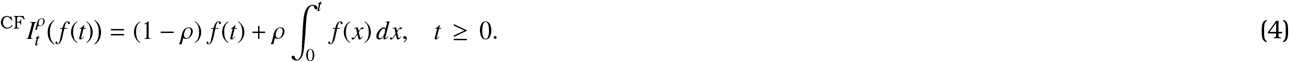

**Definition 4**. *[28] The Mittag-Leffler function is defined as:*

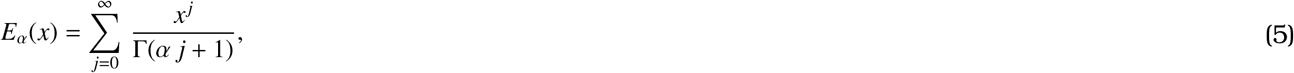

*and*

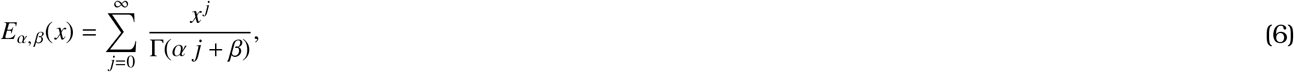

*and its Laplace transform is defined as:*

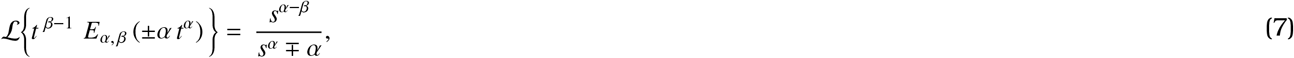

## 3. Model Formulation

Here, we consider nine disjoint classes for human population: susceptible individuals (*S* (*t*)), vaccinated individuals (*V*(*t*)), exposed individuals (*E*(*t*)), individuals infected with resistant strain *I*_*r*_(*t*), individuals infected with sensitive strain *I*_*s*_(*t*), treated individuals infected with resistant strain *T*_*r*_(*t*), treated individuals infected with sensitive strain *T*_*s*_(*t*), partially recovered individuals (carrier) *R*_*c*_(*t*) and fully recovered individuals *R*_*f*_ (*t*). Hence, the total human population, (*N*_*h*_(*t*)), can be defined as:

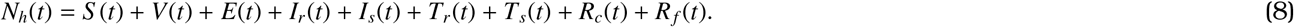

The mosquito population, *N*_*v*_(*t*), is divided into three classes: susceptible vector (*S* _*v*_(*t*)), exposed vector (*E*_*v*_(*t*)), and infected vector (*I*_*v*_(*t*)) so that

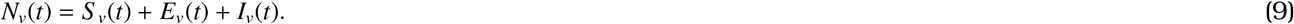

Susceptible humans get infected with malaria at a rate

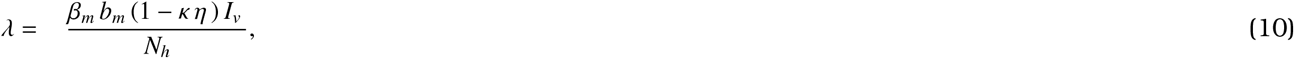

Mosquitoes become infected with malaria at a rate *λ*_*v*_, given by

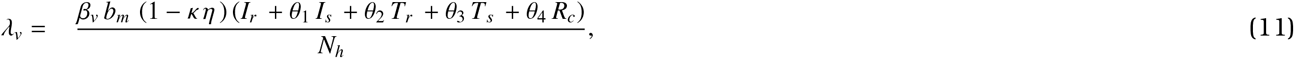

Here, *β*_*m*_ represents malaria transmission probability in humans and *b*_*m*_ represents the biting rate. *β*_*v*_ stands for the probability of malaria transmission in mosquitoes. *κ* stands for the rate of usage of malaria prevention measures (repellent, bed nets, wearing long dresses, etc.), and *η* represents the efficacy of malaria prevention measures. The modification parameters 0 *< θ*_*i*_ *<* 1, *i* = 1, 2, 3, 4, indicate low infectiousness of the individuals in the *I*_*s*_, *T*_*r*_, and *T*_*s*_ classes compared to the individuals in the *I*_*r*_ class. Λ, and Λ_*v*_ indicate the rate of inflow for humans and mosquitoes, respectively. Susceptible humans receive vaccines at a rate *ν*. However, vaccinated individuals further move to the susceptible class at a rate *α* due to the loss of vaccine-derived immunity. *λ*, (1 − *ϵ*) *λ* and *λ*_*v*_ represent the infection rate of susceptible individuals, vaccinated susceptible individuals, and susceptible mosquitoes, respectively. *ϵ* is the vaccine efficacy. *µ* refers to the natural mortality rate of humans, while *µ*_*v*_ corresponds to that of mosquitoes. Exposed humans become infected at a rate *σ*. Exposed mosquitoes move to the *I*_*v*_ class at a rate *σ*_*v*_. We have considered two classes of infected individuals: individuals infected with drug-resistant strains (*I*_*r*_) and Individuals infected with drug-sensitive strains (*I*_*s*_). Exposed individuals move to *I*_*r*_ and *I*_*s*_ classes at a rate *b σ* and (1 − *b*) *σ*, respectively, where *b* is a modification parameter. Individuals in the *I*_*r*_ class receive treatment at a rate *τ*_*r*_. Individuals in the *I*_*s*_ class receive treatment at a rate *τ*_*s*_. We have also considered two classes for recovered individuals: fully recovered individuals (*R*_*f*_) and individuals who recover from the disease but carry the parasite (*R*_*c*_). It is assumed that due to natural immunity individuals in the *I*_*r*_, *I*_*s*_ classes recover from malaria but always carry the parasite. Individuals in the *I*_*r*_, *I*_*s*_, *T*_*r*_, *T*_*s*_ classes progress to *R*_*c*_ class at a rate *ψ*_*irc*_, *ψ*_*isc*_, *ψ*_*trc*_, and *ψ*_*tsc*_, respectively. Individuals in the *T*_*r*_ and *T*_*s*_ classes recover at a rate *ψ*_*tr f*_, and *ψ*_*ts f*_, respectively. Disease-induced death rate for individuals in the *I*_*r*_, *I*_*s*_, *T*_*r*_, and *T*_*s*_ classes are *δ*_*ir*_, *δ*_*is*_, *δ*_*tr*_, and *δ*_*ts*_, respectively. *ξ*_*c*_ and *ξ* _*f*_ represent the re-infection rate of individuals from *R*_*c*_ and *R*_*f*_ classes, respectively.

Based on the above discussion, we derive the following system of differential equations to capture the dynamics of malaria. Therefore, the system of differential equations to model the malaria transmission dynamics is as follows:

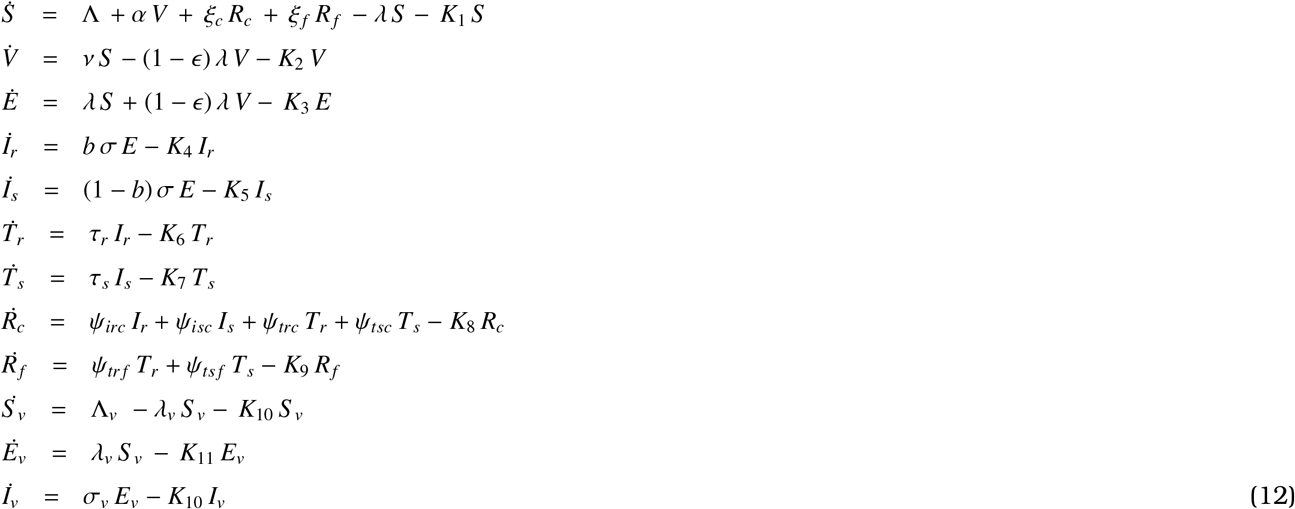

where,

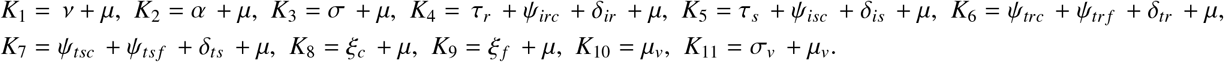

**Figure 1.**
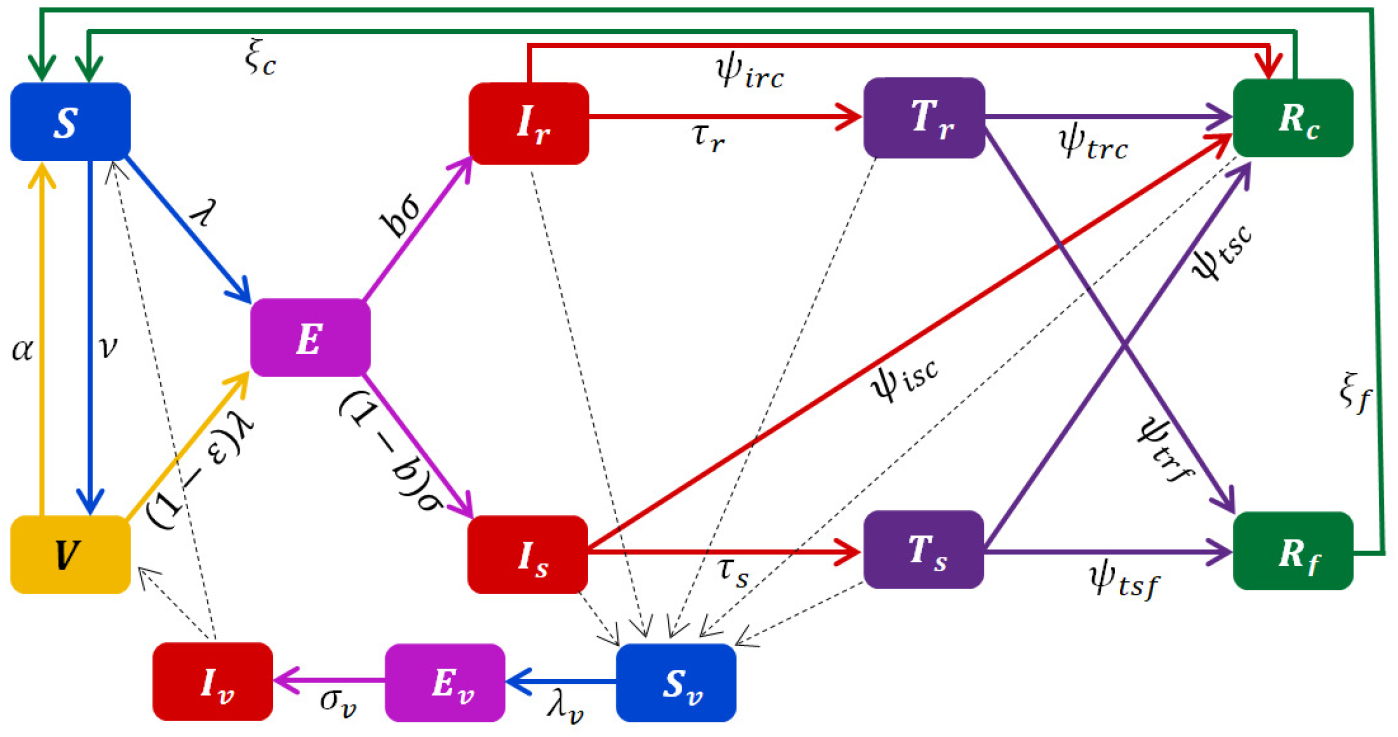
Diagram shows malaria transmission as described in system (12).

As the classical integer order derivative is local in nature and does not provide any memory effect, we have modified model (12) to the following CFFOD model as follows:

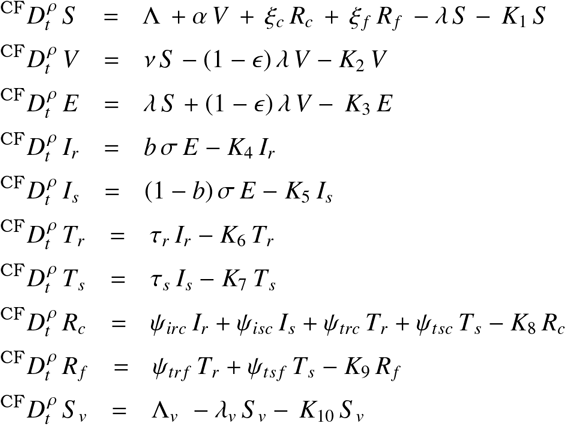

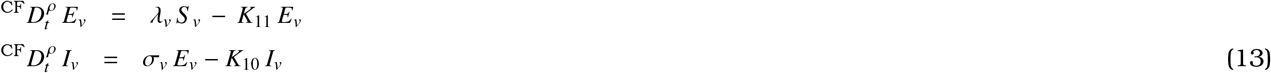

with

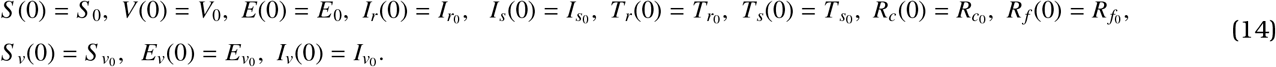

## 4. Theoretical analysis of the model

### 4.1 Positivity and boundedness

considering all the equations of system (13) and using (8), we obtain

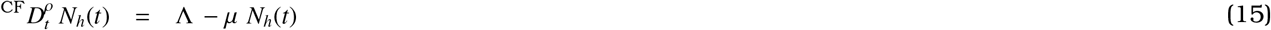

Applying Laplace transformation on both sides of equation (15) we have,

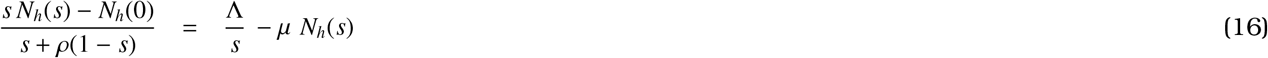

Further, equation (16) simplifies to

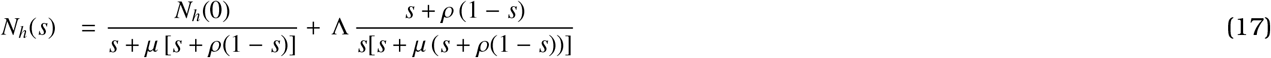

Applying inverse Laplace transformation on (17) and performing some calculations, we get,

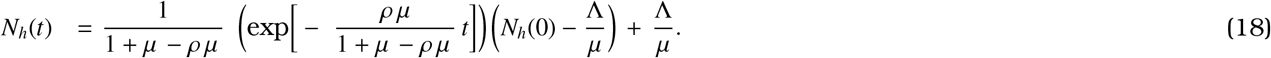

If 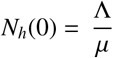, then 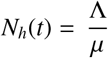. Also if 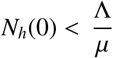, then 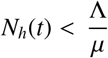.

Thus 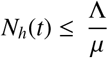.

For the positivity of the system solutions,

Let, 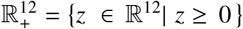 and *z*(*t*) {*S* (*t*), *V*(*t*), *E*(*t*), *I*_*r*_ (*t*), *I*_*s*_ (*t*), *T*_*r*_ (*t*),*T* _*s*_ (*t*), *R*_*c*_ (*t*), *R*_*f*_ (*t*), *S*_*v*_ (*t*), *E*_*v*_ (*t*), *I*_*v*_ (*t*)}^*T*^

Thus, we have the following corollary [29].

**Corollary 1**. *Suppose that, g*(*t*) ∈ *CF*[*m, n*] *and* 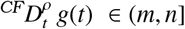 *where, ρ* ∈ (0, 1]. *If*

(*a*) 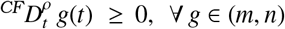, *then g*(*t*) *is non-decreasing*.

(*b*) 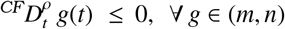, *then g*(*t*) *is non-increasing*.

Now, we claim the following proposition.

**Proposition 1**. *The solution of the model* (13) - (14) *is non-negative and bounded for all* 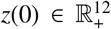 *for t >* 0.

*Proof*. From system (13), we have,

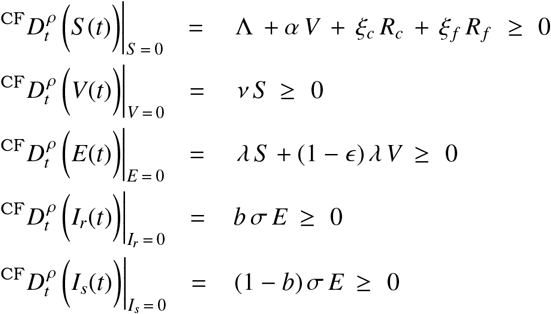

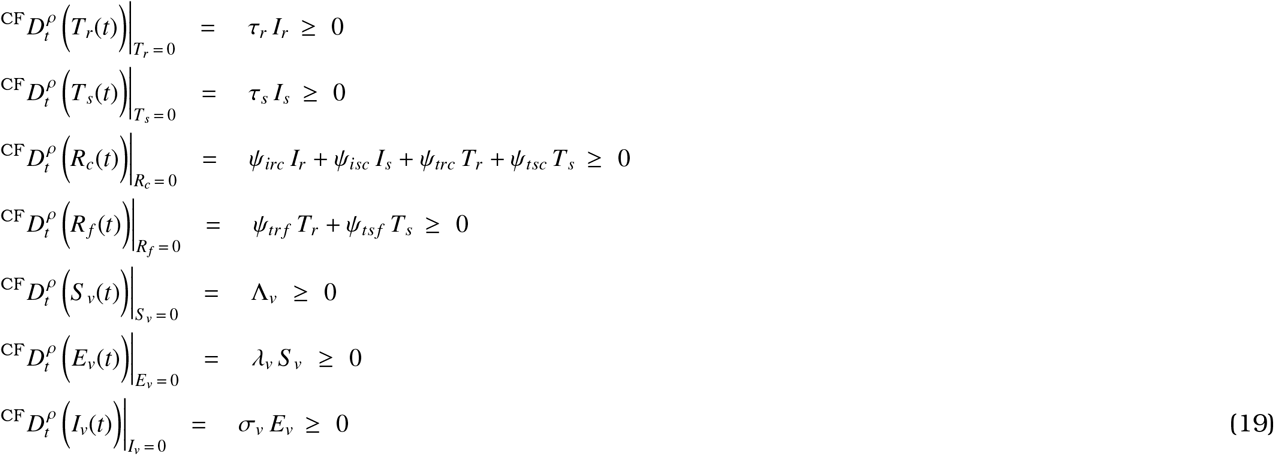

Using the above corollary, it is therefore concluded that any solutions

*S* (*t*), *V*(*t*), *E*(*t*), *I*_*r*_(*t*), *I*_*s*_(*t*), *T*_*r*_(*t*), *T*_*s*_(*t*), *R*_*c*_(*t*), *R*_*f*_ (*t*), *S* _*v*_(*t*), *E*_*v*_(*t*), *I*_*v*_(*t*) of model (13) with initial conditions (14) are non-negative for all *t >* 0.

### 4.2 Local stability of the disease free equilibrium (DFE)

Here, we start with computing the control reproduction number (*ℛ*_*m*_). For this, we follow the procedure discussed in [30, 31]. Using the notation of [31], from the system (12) we have

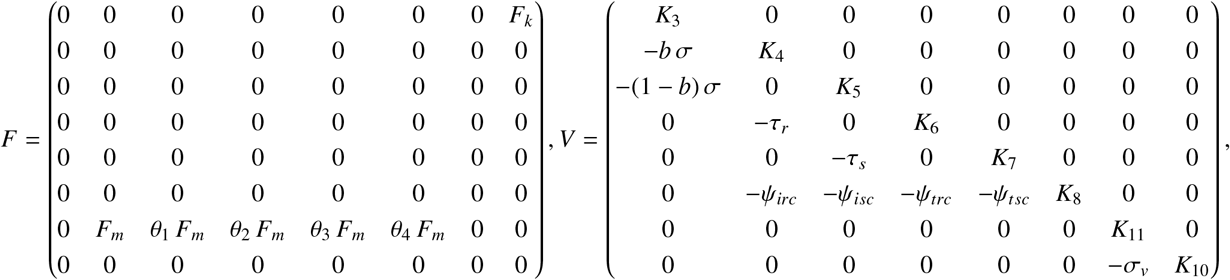

Following the method from [32, 33], the expression of *ℛ* _*m*_, can be calculated as

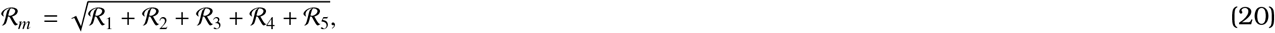

where,

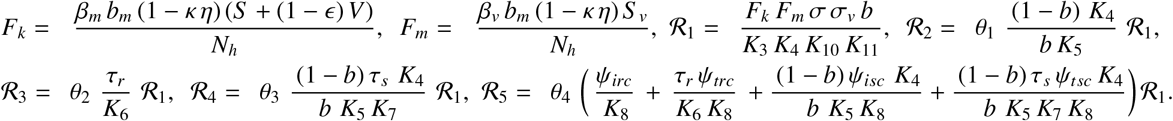

Now, the following can be stated based on Theorem 2 of [31].

**Lemma 1**. *The DFE* (E_0_) *of the model* (12) *is locally-asymptotically stable (LAS) if ℛ*_*m*_ *<* 1 *and unstable if ℛ*_*m*_ *>* 1.

*Proof*. To prove this, we first calculate the Jacobian of the system (13) at the DFE which is given as

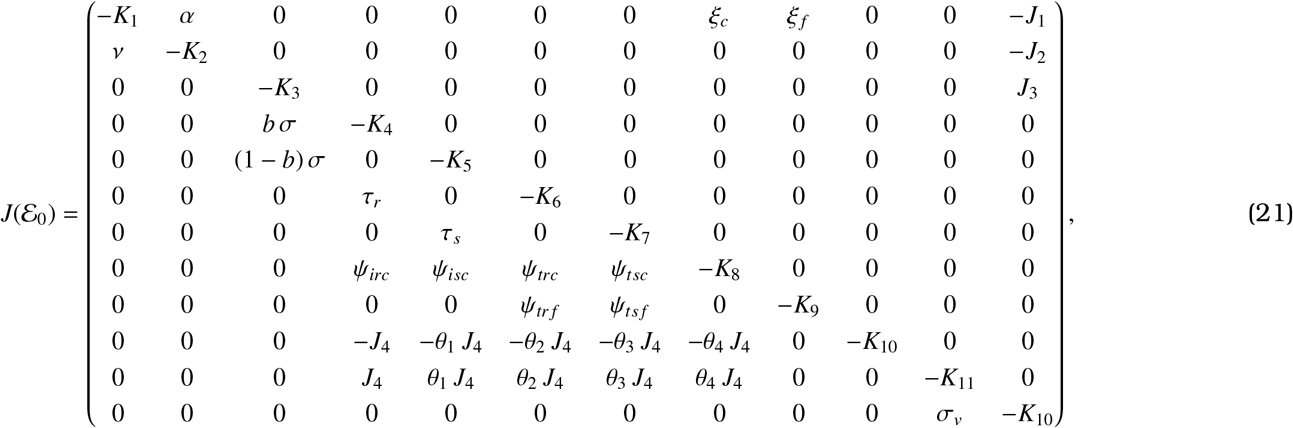

where,

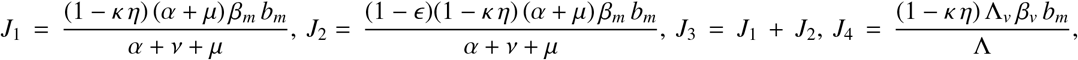

Now, according to [31], the control reproduction number (*ℛ*_*m*_) will be less than one if the eigenvalues of the matrix (21) are either negative or the real part of them are negative. Now, this is possible if the following condition is satisfied.

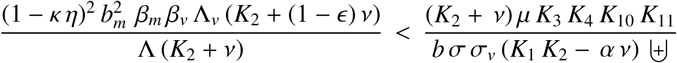

where, 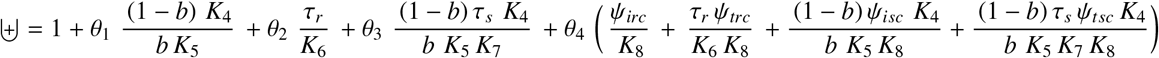,Consequently, this condition leads to the conclusion that *ℛ*_*m*_ *<* 1.

### 4.3 Sensitivity analysis of the model parameters

To determine the parameters that significantly influence the reproduction number, we employ the normalized forward sensitivity index method, as described in [34, 35, 36]. The sensitivity index of the reproduction number *ℛ*_*m*_ with respect to a parameter *ω* is defined as

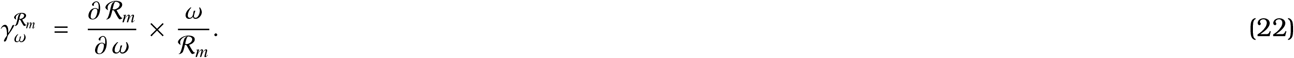

Using formula (22), for some of the model parameters, we have:

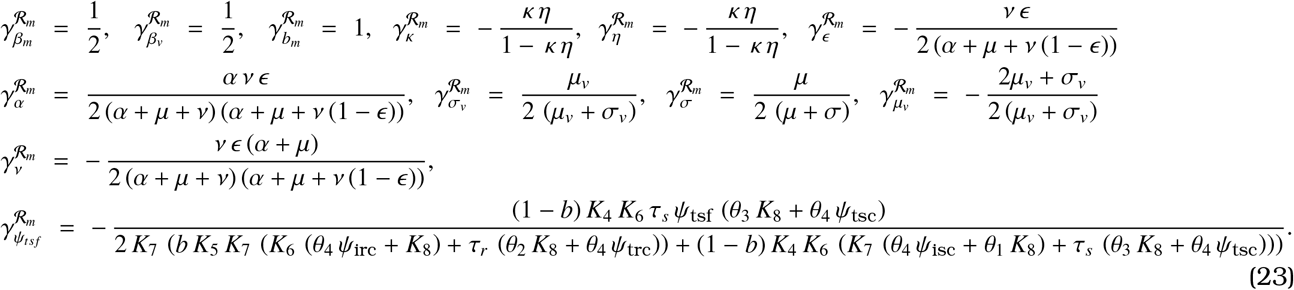

A positive value of sensitive index corresponding to a parameter implies that if the parameter values is increased (decreased), *ℛ*_*m*_ will increase (decrease). A negative value of sensitive index represents an inverse relationship between *ℛ*_*m*_ and parameter values.

As, all the parameters are positive and 0 *< ϵ, b, κ η <* 1, parameters *β*_*m*_, *β*_*v*_, *b*_*m*_, *α, σ, σ*_*v*_ are positively correlated to *ℛ*_*m*_ and hence, *ℛ*_*m*_ can be brought under one if these parameter values are reduced accordingly. However, parameters *κ, η, ϵ, µ*_*v*_, *ν, ψ*_*ts f*_ have negative influence on *ℛ*_*m*_ and hence, *ℛ*_*m*_ can be brought under unity by properly increasing values of these parameters.

### 4.4 Existence and uniqueness

Applying formula (2) to system (13), we obtain,

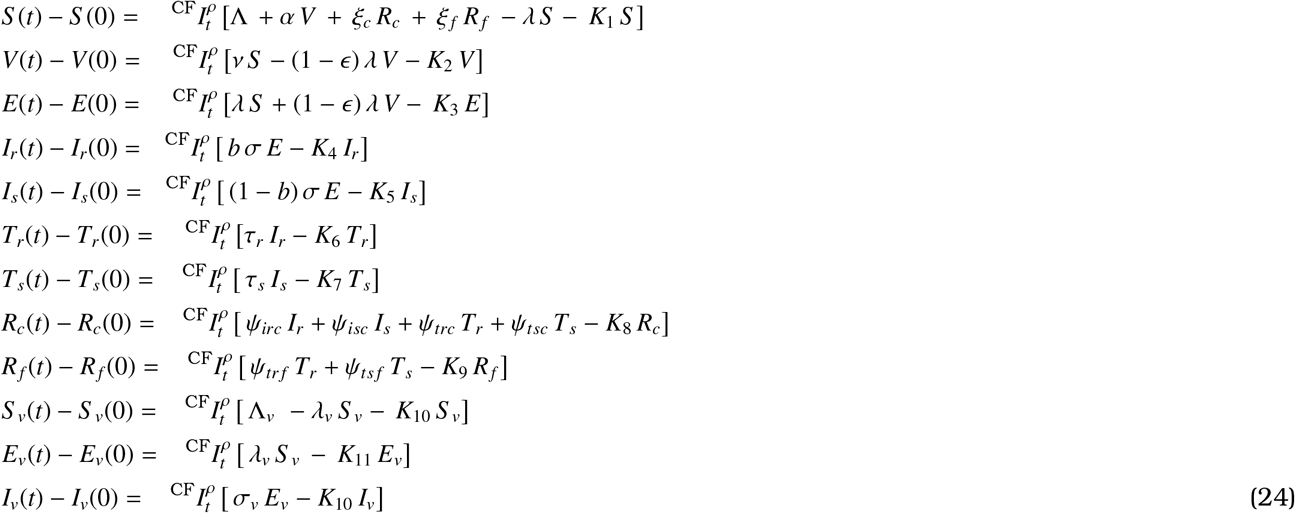

For convenience, we define the following kernels

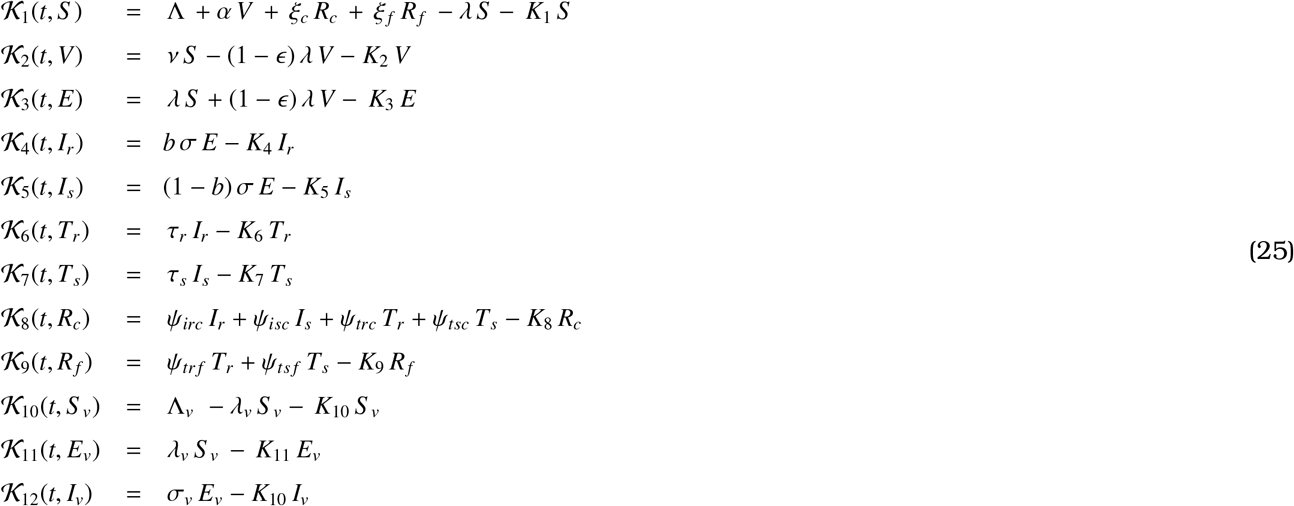

and the function

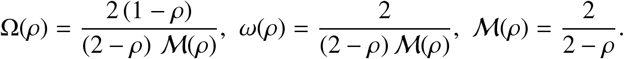

To prove Theorem 1, we assume that *S* (*t*), *V*(*t*), *E*(*t*), *I*_*r*_(*t*), *I*_*s*_(*t*), *T*_*r*_(*t*), *T*_*s*_(*t*), *R*_*c*_(*t*), *R*_*f*_ (*t*), *S* _*v*_(*t*), *E*_*v*_(*t*), *I*_*v*_(*t*) are non-negative bounded function, i.e. ∥*S* (*t*)∥ ≤ Ξ_1_, ∥*V*(*t*)∥ ≤ Ξ_2_, ∥*E*(*t*)∥ ≤ Ξ_3_, ∥*I*_*r*_(*t*)∥ ≤ Ξ_4_, ∥*I*_*s*_(*t*)∥ ≤ Ξ_5_, ∥*T*_*r*_(*t*)∥ ≤ Ξ_6_, ∥*T*_*s*_(*t*)∥ ≤ Ξ_7_, ∥*R*_*c*_(*t*)∥ ≤ Ξ_8_, ∥*R*_*f*_ (*t*)∥ ≤ Ξ_9_, ∥*S* _*v*_(*t*)∥ ≤ Ξ_10_, ∥*E*_*v*_(*t*)∥ ≤ Ξ_11_, ∥*I*_*v*_(*t*)∥ ≤ Ξ_12_.

Applying Definition 2 to (24), we obtain

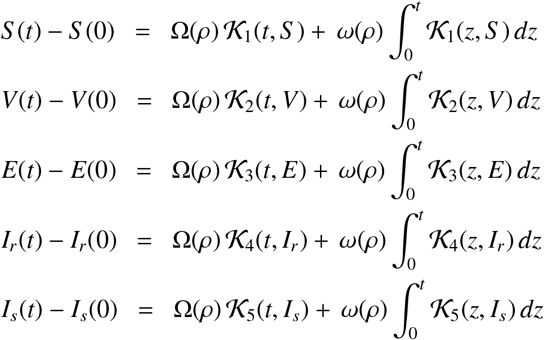

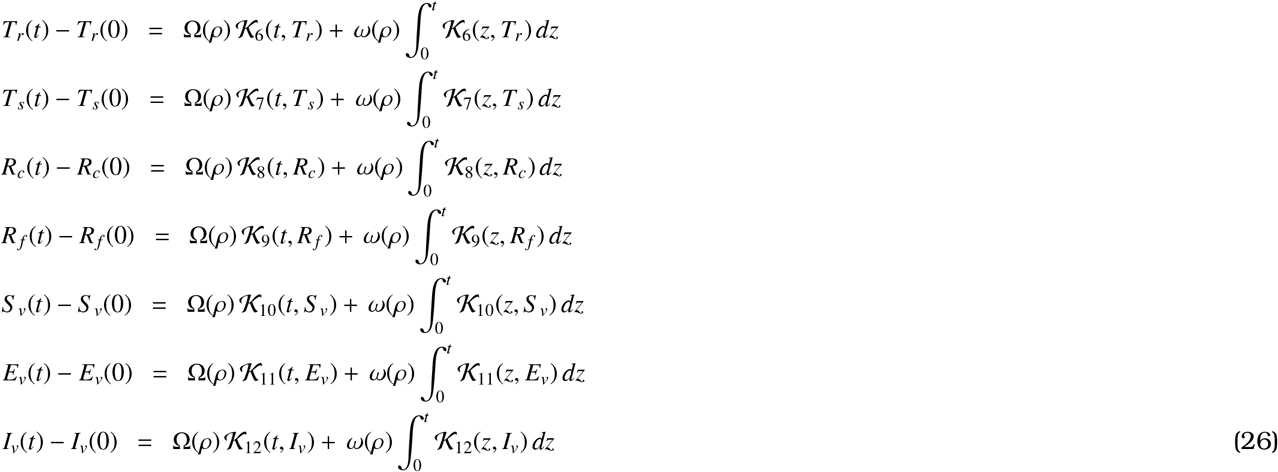

**Theorem 1**. *The functions 𝒦*_*i*_, *for i* = 1, 2, …, 12, *satisfy Lipshitz’s conditions and are contraction mappings, if the following condition holds*

0 ≤*ℳ*^*^ = *max*{ℵ_1_, ℵ_2_, ℵ_3_, ℵ_4_, ℵ_5_, ℵ_6_, ℵ_7_, ℵ_8_, ℵ_9_, ℵ_10_, ℵ_11_, ℵ_12_} *<* 1.

*Proof*.

Let us consider the kernel *𝒦* _1_. For any two functions *S* and *S* _1_, we have

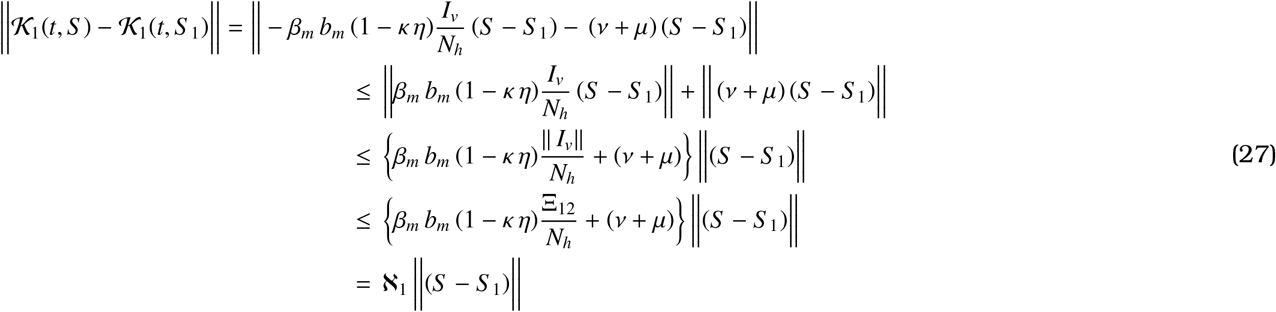

where,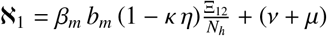.

Thus *𝒦*_1_ satisfies the Lipschitz condition. Similarly, it is possible to find ℵ_*i*_, *i* = 2, 3, …, 12 for which the Lipschitz’s conditions are satisfied by *𝒦*_*i*_, *i* = 2, 3, …, 12. Moreover, under the condition, 0 ≤*ℳ*^*^ = max {ℵ_1_, ℵ_2_, ℵ_3_, ℵ_4_, ℵ_5_, ℵ_6_, ℵ_7_, ℵ_8_, ℵ_9_, ℵ_10_, ℵ_11_, ℵ_12_}*<* 1 the functions are contractions. Again, equation (26) can be written in terms of the kernels as follows:

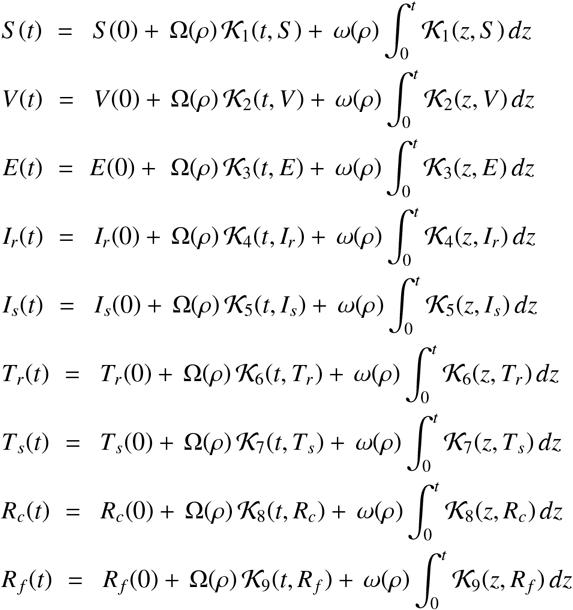

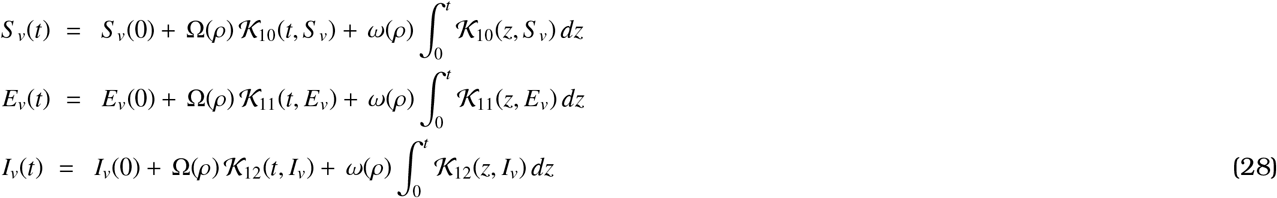

From equation (28), we can have the following recursive relations:

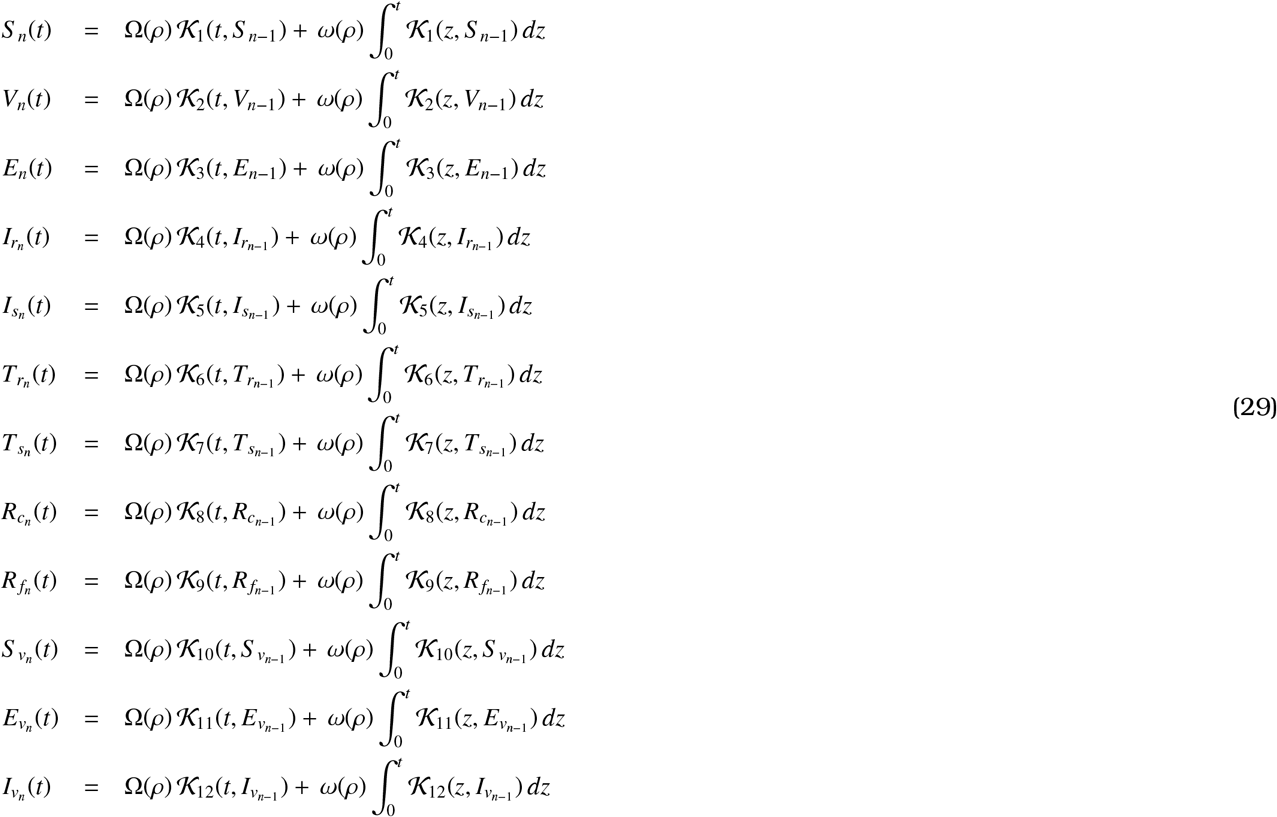

Now, we have

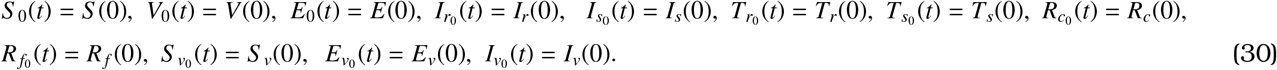

Again, we can write

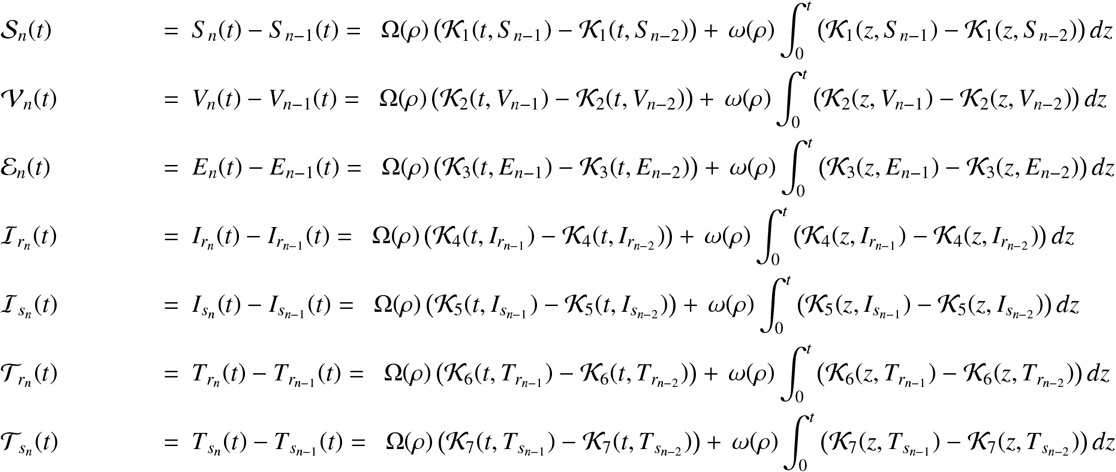

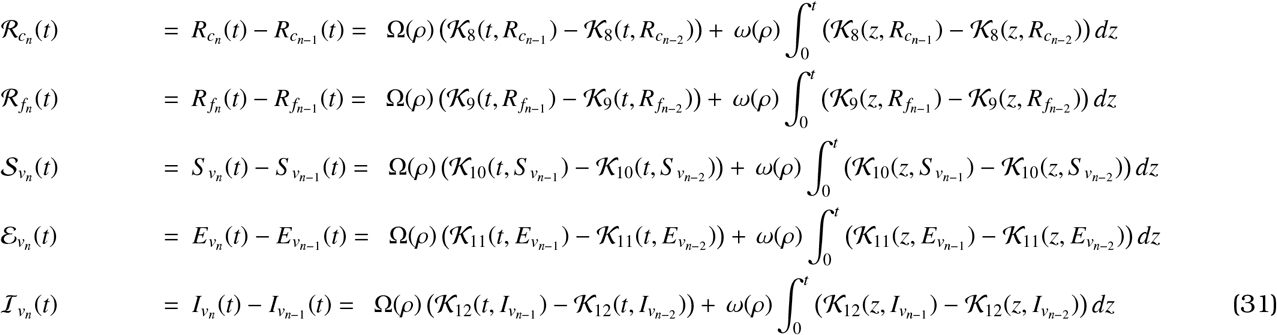

where,

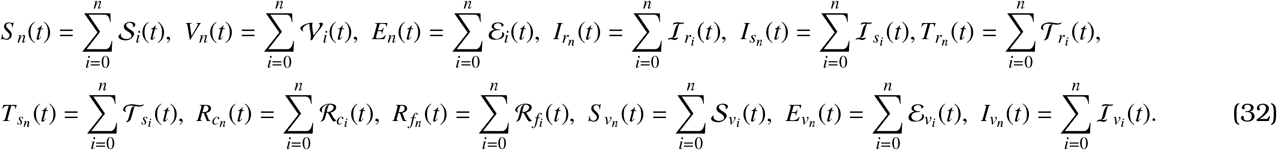

Next, we have

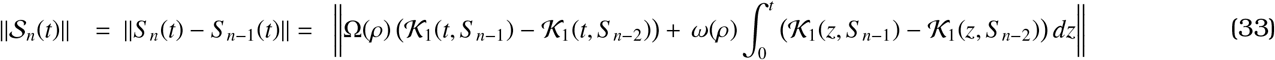

Using the triangle inequality from equation (33), the following can be obtained,

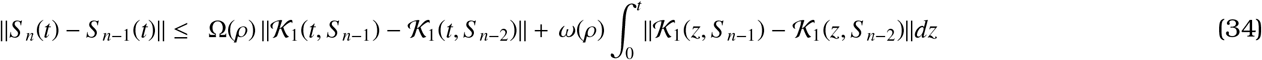

As the kernal *𝒦*_1_ satisfies the Lipschitz condition with ℵ_1_ as the Lipschitz constant, we have from equation (34)

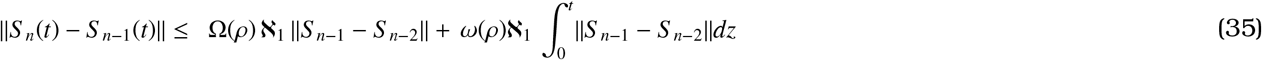

Thus, we obtain,

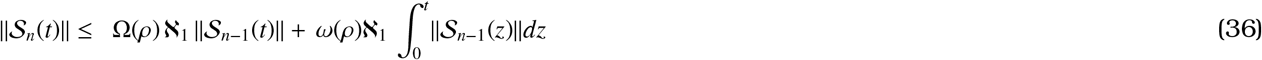

Similarly,

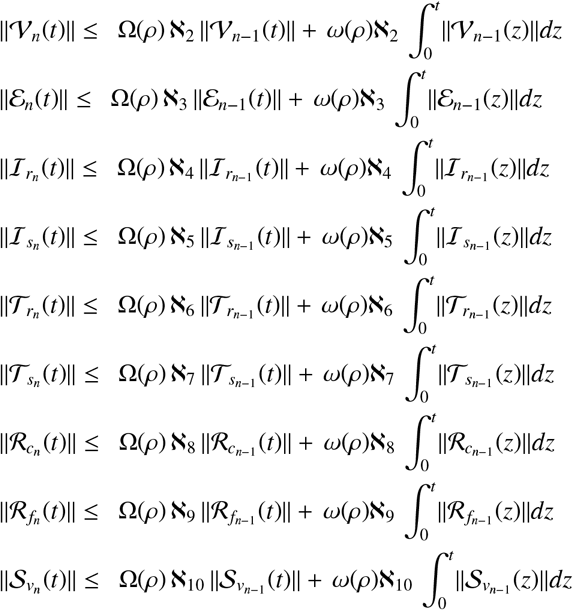

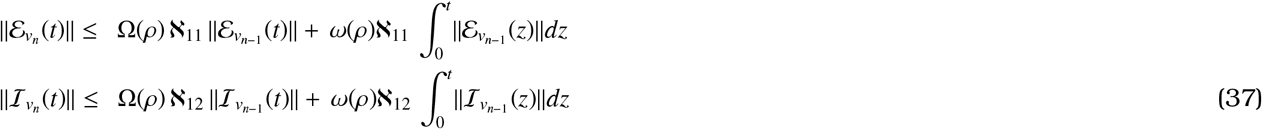

**Theorem 2**. *If the inequality* Ω(*ρ*) ℵ_*i*_ + *ω*(*ρ*) ℵ_*i*_ *t*_0_ *<* 1, *i* = 1, 2, …, 12 *holds for any time t*_0_ *>* 0 *then a system of solution exists for the model* (13) *with initial condition* (14).

*Proof*.

Since all of *S* (*t*), *V*(*t*), *E*(*t*), *I*_*r*_(*t*), *I*_*s*_(*t*), *T*_*r*_(*t*), *T*_*s*_(*t*), *R*_*c*_(*t*), *R*_*f*_ (*t*), *S* _*v*_(*t*), *E*_*v*_(*t*), and *I*_*v*_(*t*) are bounded and each of the *𝒦*_*i*_, *i* = 1, 2, …, 12 satisfies a Lipschitz condition, using equations (36) and (37), we have the following:

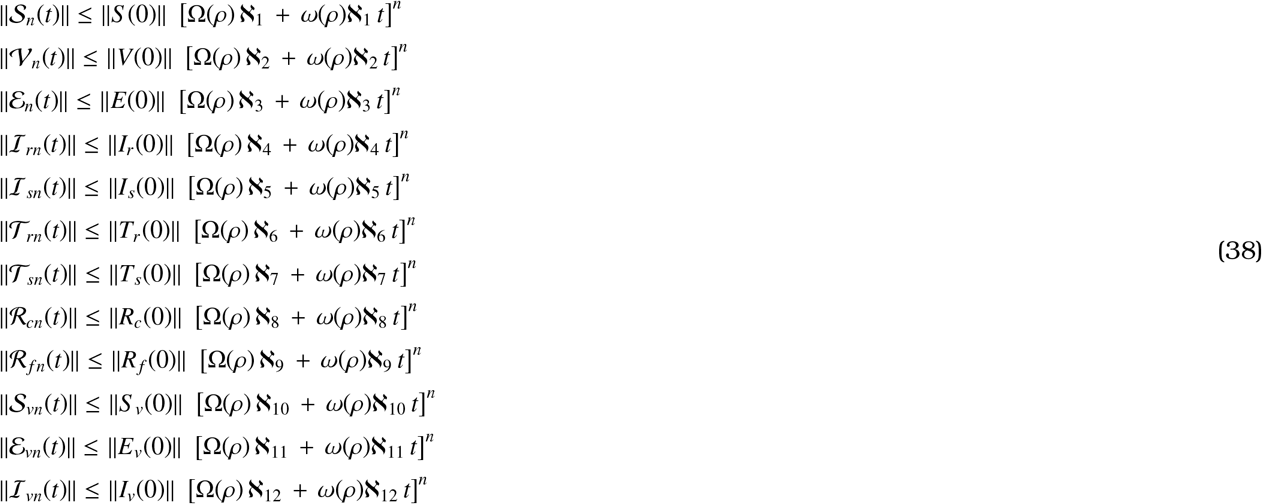

Equation (38) implies that there exist functions defined in (32). It also ensures that the functions are smooth. Now it is sufficient to show that 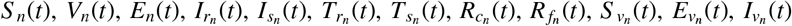 converge to a solution of (13)-(14). Now, the remainder terms after *n* iterations for each class can be defined as Σ_1*n*_(*t*), Σ_2*n*_(*t*), Σ_3*n*_(*t*), Σ_4*n*_(*t*), Σ_5*n*_(*t*), Σ_6*n*_(*t*), Σ_7*n*_(*t*), Σ_8*n*_(*t*), Σ_9*n*_(*t*), Σ_10*n*_(*t*), Σ_11*n*_(*t*), Σ_12*n*_(*t*), i.e.

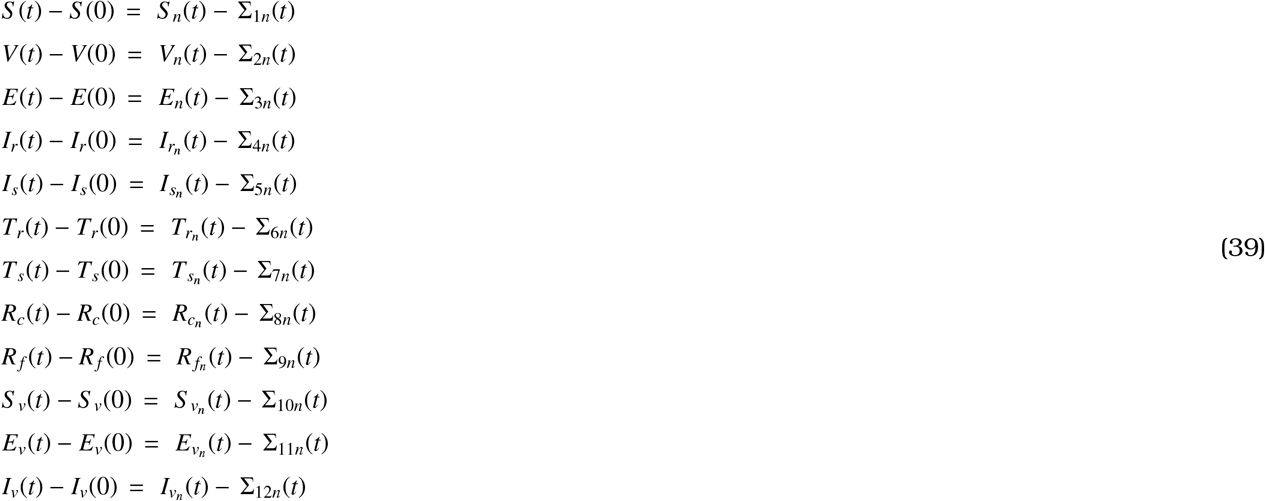

Hence, we have

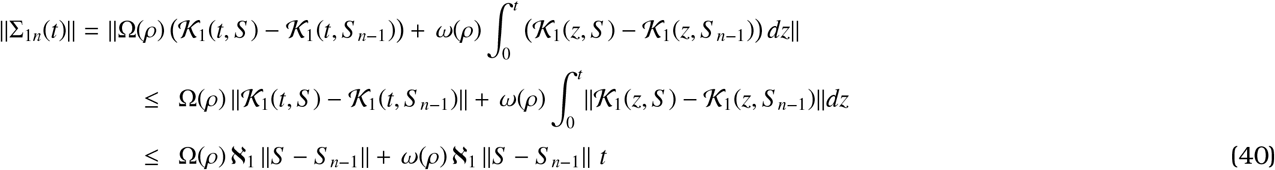

After some recursive process, we obtain,

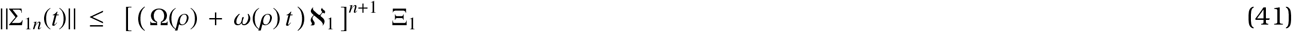

Then at *t*_0_, we obtain

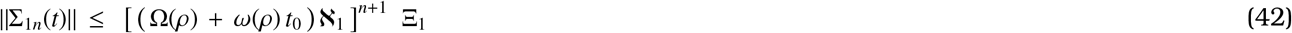

Taking limit on equation (42) as *n* → ∞, we get,

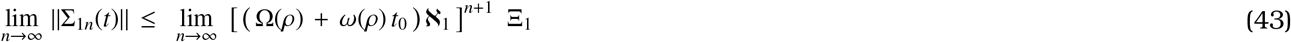

Using hypothesis Ω(*ρ*) ℵ_1_ + *ω*(*ρ*) ℵ_1_ *t*_0_ *<* 1 equation (43) becomes,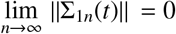.

Similarly, we obtain,

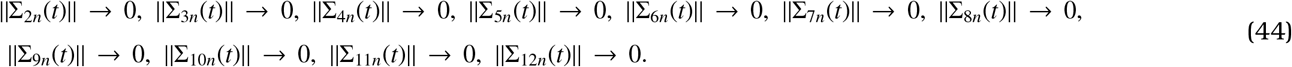

Therefore, Theorem 2 is proved.

Now, to prove the uniqueness of the solutions we begin with proving Theorem 3.

**Theorem 3**. *The system* (13) *with* (14) *has unique solutions if the following conditions hold:*

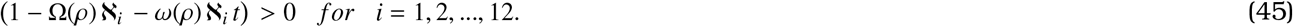

*Proof*.

First suppose that 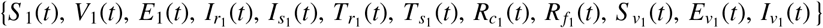 is another solution set of the model (13)-(14). Then, it can be written

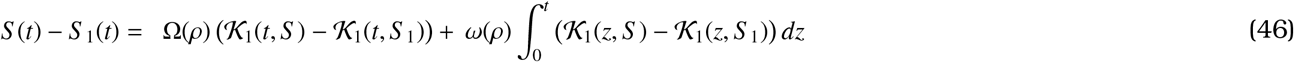

Applying norm to the equation (46) the following can be obtained.

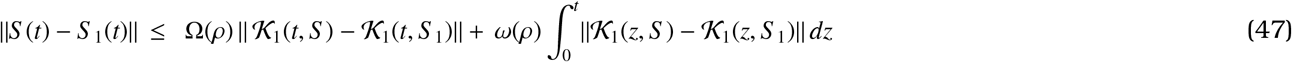

Using the Lipschitz condition (47) becomes

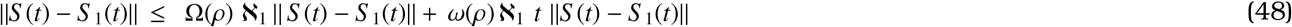

This implies

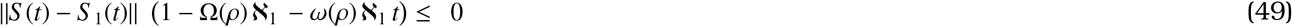

Now, with condition (45), equation (49) becomes

∥*S* (*t*) − *S* _1_(*t*)∥ = 0.

Thus, *S* (*t*) = *S* _1_(*t*).

Similarly, using inequality (45), it can be shown that

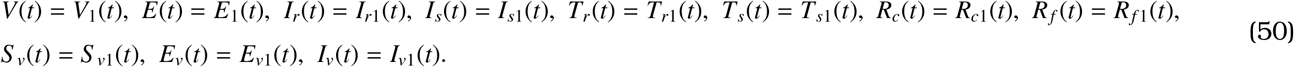

This completes the proof.

## 5. Three-step Adams–Bashforth Numerical scheme

To perform numerical simulation of the model (12), we use three-step Adams–Bashforth method. We will use the procedure described in [27]. Consider the following CFFDE

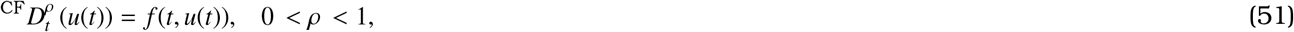

where 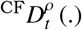 represents CFFOD. Now, using the procedure described in [27], we have

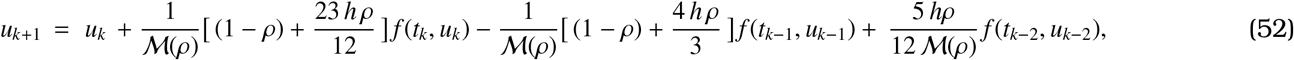

where *u*_*k*+1_ = *u*(*t*_*k*+1_), *u*_*k*_ = *u*(*t*_*k*_), *u*_*k*−1_ = *u*(*t*_*k*−1_), *u*_*k*−2_ = *u*(*t*_*k*−2_), and *u*_0_ = *u*(*t*_0_) = [*S* (*t*_0_), *V*(*t*_0_), *E*(*t*_0_), *I*_*r*_(*t*_0_), *I*_*s*_(*t*_0_), *T*_*r*_(*t*_0_), *T*_*s*_(*t*_0_), *R*_*c*_(*t*_0_), *R*_*f*_ (*t*_0_), *S* _*v*_(*t*_0_), *E*_*v*_(*t*_0_), *I*_*v*_(*t*_0_)]^*T*^.

## 6. Optimal Control

To prevent the disease spread, reduce the disease related complexity, and minimize the mortality, we introduce three control measures (*u*_1_, *u*_2_, and *u*_3_) in our existing model. Control variable *u*_1_ is introduced to prevent individuals from being infected with malaria. This control refers that people are warned through advertisements about how to protect themselves from mosquitoes, destroying mosquito breeding grounds, and the devastating socio-economic effects of malaria. Control measure *u*_2_ is initiated to ensure the continuous supply of malaria vaccines. Control measure *u*_3_ refers that people are getting proper treatment and medical care against malaria. Incorporating all these features, the optimal control model of the basic malaria model (12) can be formulated as follows:

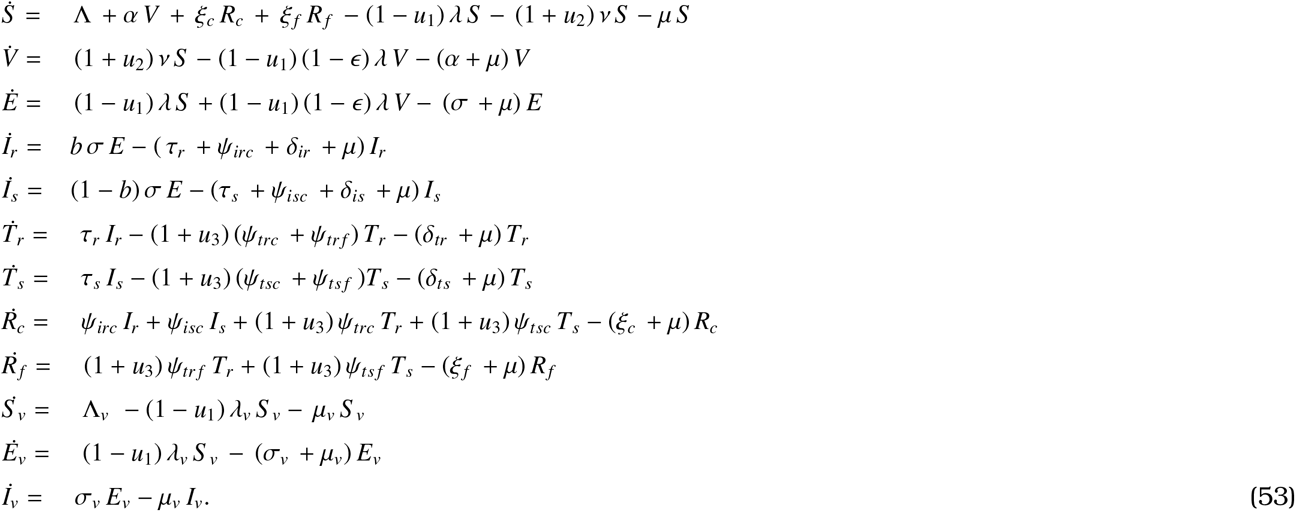

The objective of this section is to find appropriate control strategies to minimize the infected population and the implementation costs of the control strategies. We also concentrate on achieving the maximum outcomes through the implementation of control measures. In this study, we have considered quadratic objective functional as the quadratic functionals are differentiable and convex [37]. Also, convexity ensures the existence of a unique optimal control [38]. Furthermore, quadratic costs align with real-world situations as cost increases non-linearly. For all these, we have formulated the following objective functional (equation (54)) [37, 38, 39].

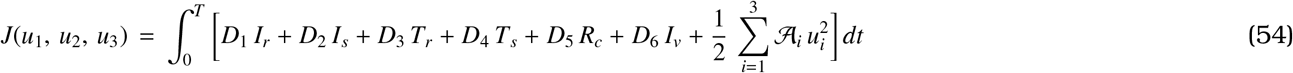

where, *D*_*i*_, and *𝒜* _*j*_, *i* = 1, 2, ….., 6 and *j* = 1, 2, 3 are the balancing cost factors.

**Theorem 4**. *Assume that control functions defined above* (53) *are Lebesgue integrable and bounded on* 0 ≤ *t* ≤ *T*. *Then we have an optimal control triplet* 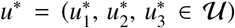 *such that*

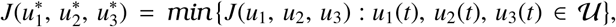

*where, 𝒰 is the control set. We define 𝒰 as*

*𝒰* = (*u*_1_, *u*_2_, *u*_3_) : *u*_*i*_(*t*) *is measurable on* [0, *T*], 0 ≤ *u*_*i*_(*t*) ≤ 1, *i* = 1, 2, 3.

This can be proved as in [37].

### 6.1 Characterization of the optimality system and the Hamiltonian

To obtain the expression of the Hamiltonian (H), we will apply the Pontryagin’s maximum principle [40] to the system (53) and (54).

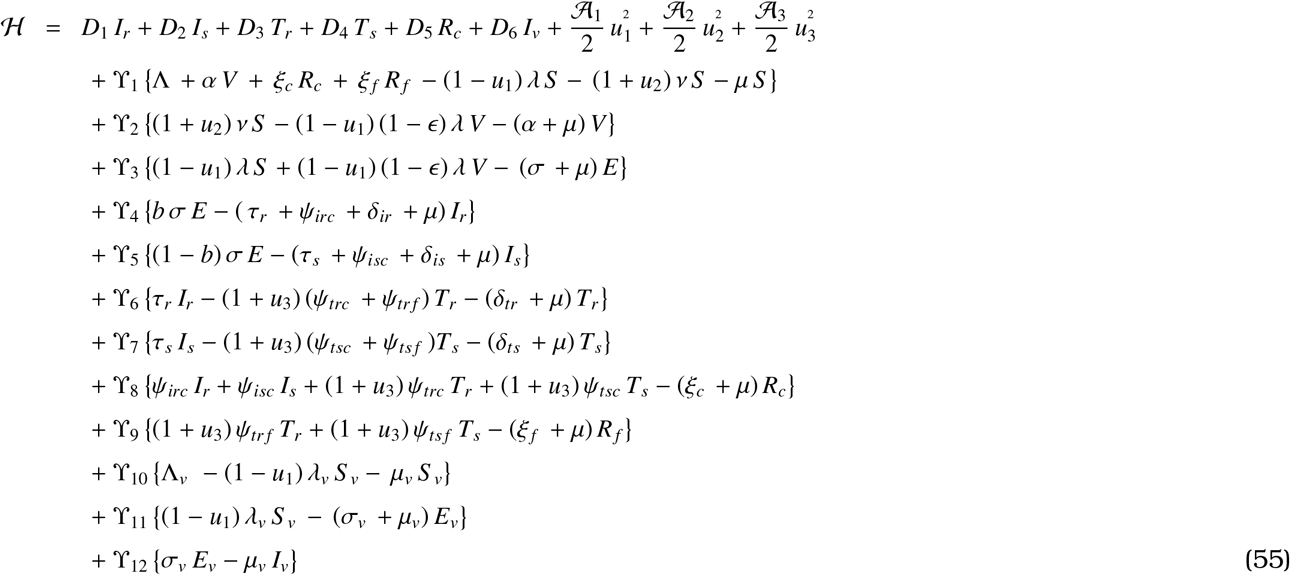

where *Υ*_*i*_, *i* = 1, 2⃜,12 are the adjoint variables.

**Theorem 5**. *If* 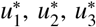 *are given control variables and S* _1_ = *S, S* _2_ = *V, S* _3_ = *E, S* _4_ = *I*_*r*_, *S* _5_ = *I*_*s*_, *S* _6_ = *T*_*r*_, *S* _7_ = *T*_*s*_, *S* _8_ = *R*_*c*_, *S* _9_ = *R*_*f*_, *S* _10_ = *S* _*v*_, *S* _11_ = *E*_*v*_, *S* _12_ = *I*_*v*_ *are solutions of the system* (53), *then adjoint variables, Υ*_*i*_, *i* = 1, 2,, 12 *can be obtained from*

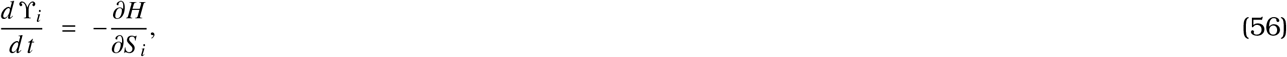

*with Υ*_*i*_(*T*) = 0. *Furthermore, we have the controls*, 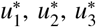, *given by*

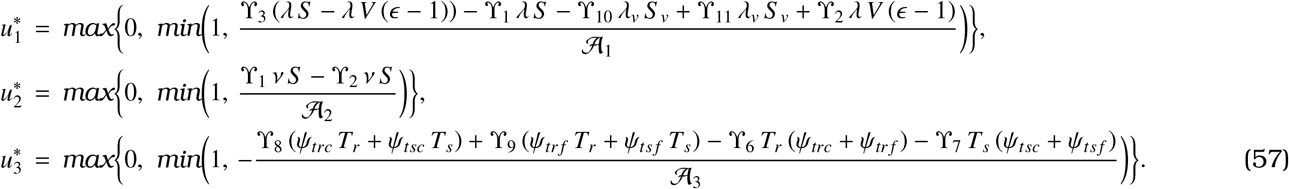

**Proof:** The proof of the existence of an optimal control follows directly from the results in [41], and is therefore omitted here. The differential equations containing the adjoint variables are achieved by differentiating the Hamiltonian with respect to the state variables.

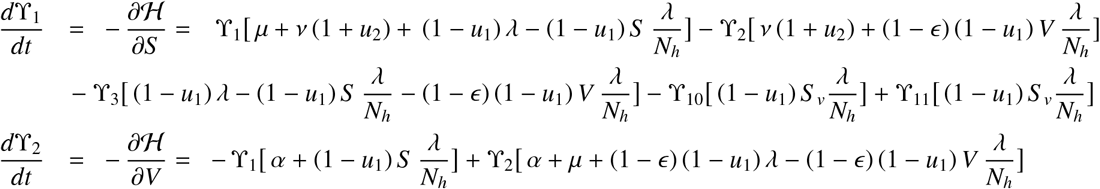

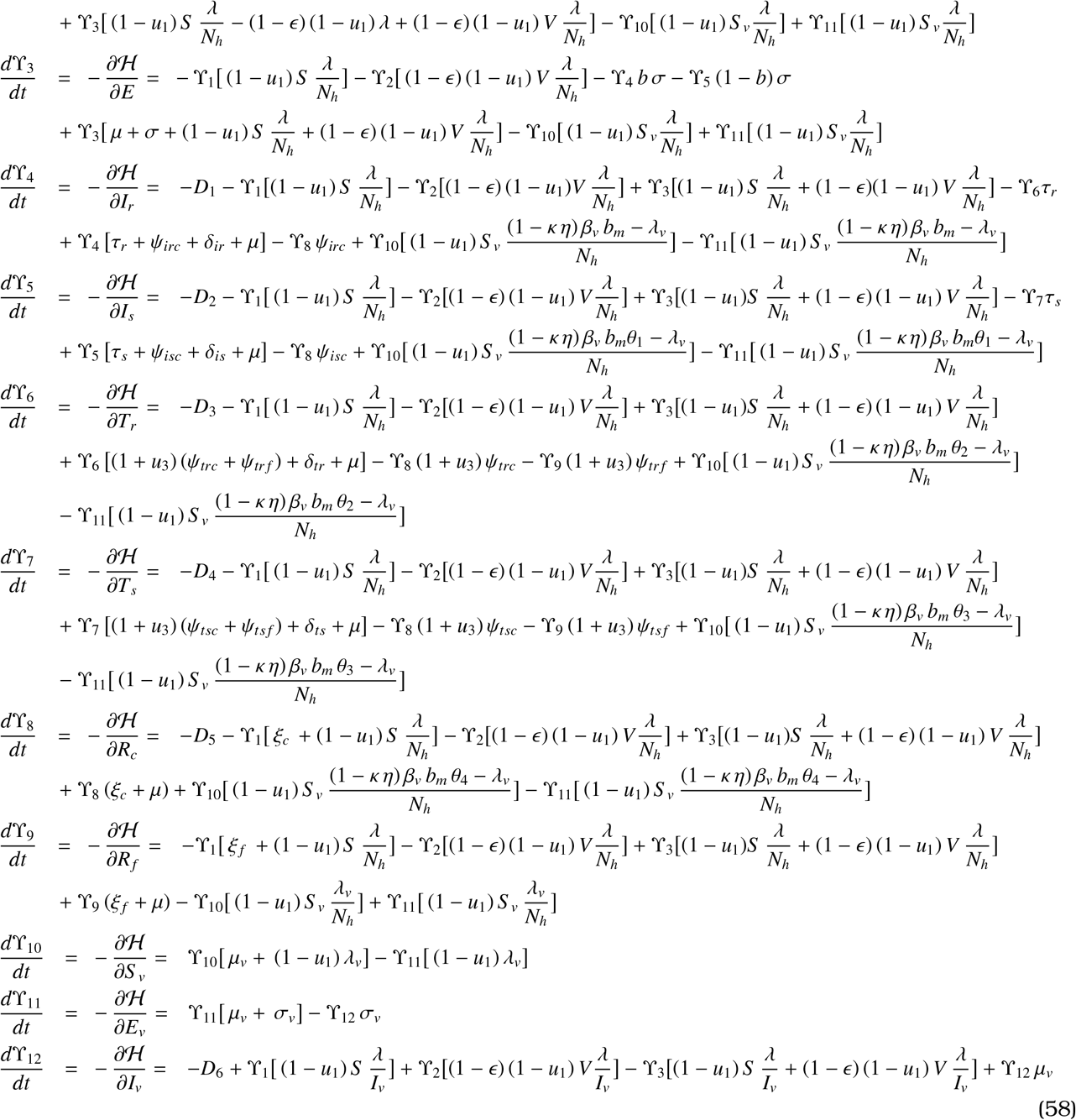

Again calculating 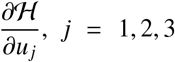, and solving the system 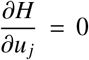 we get,

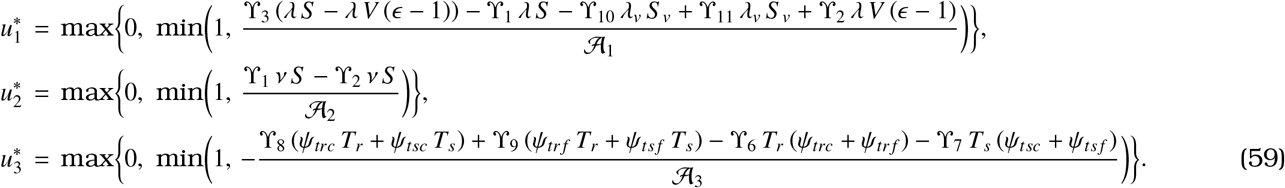

## 7. Numerical simulations

Numerical simulations are performed using Matlab to generate graphical results for the model (13) with values from Table 1. At first we compare the real data of 22 years (2000 − 2021) obtained from the World Bank [44] for Nigeria with the proposed model to provide information about the plausibility of the model. Computational works are also carried out to determine the impact of drug-resistant strains and recovered carriers on the disease dynamics. We have also analyzed how basic reproduction number behaves when some parameter values are varied. Accordingly, numerical investigations are performed to determine the effect of those parameters on the number of infected cases by plotting the infected classes by varying some of those parameter values. In Figure 2, the simulated result of the model (13) is compared with the real data. A goodness of fit test (using normalized root mean squared error, NRMSE) is also performed to show how closely calculated data mirrors the real data. If the fit value is 0, this indicates a perfect fit between the estimated data and the real data. In our case, the fit value is 0.1012 which shows a close connection between the real data and the simulated result. Numerical Simulation is also carried out to show the stability of the model solution when the reproduction number (*ℛ*_*m*_) is either less than one or greater than one. Figure 3 shows that solution trajectories tend to DFE when *ℛ*_*m*_ *<* 1 which verifies the theoretical result. However, Figure 4 depicts that solution trajectories tend to EEP when *ℛ*_*m*_ *>* 1. Further, Figure 5 shows that solution trajectories tend to both DFE and EEP even when *ℛ*_*m*_ *<* 1 with Λ = 215000, Λ_*v*_ = 20000, *κ* = 0.009, *ξ* _*f*_ = 0.2, *ξ*_*c*_ = 0.2, *ν* = 10, *α* = 10 and other parameter values as in Table 1. This implies the possibility of the occurrence of backward bifurcation. In Figures 6 (a) - (f), approximate solutions of different compartments (infected, treatment, and recovered) are presented by changing the values of order (*ρ*). These figures show the results for *ρ* = 0.9, 0.95, 1.0 and according to these figures, the rate of decay and growth depends on the order of the fraction. Figures 6 (a) (f) show that the larger the order the slower the process and vice versa. In the rest of the simulations, we have considered *ρ* = 0.95. In Figure 7, the effect of *β*_*m*_ and *β*_*v*_ on *ℛ*_*m*_ and infected cases are presented. Figures 7 (a) and (c) show that R_*m*_ increases with an increase in *β*_*m*_ and *β*_*v*_. A similar trend is observed for the infected cases (Figures 7 (b) and (d)). For instance, Figures 7 (b) and (d) demonstrate that there is roughly 40 % and 75 % increase in the peak of daily infected patients when *β*_*m*_ increases from 0.35 to 0.55 and *β*_*v*_ increases from 0.1 to 0.3, respectively. We also conduct simulation to investigate how *b*_*m*_ and *µ*_*v*_ influence the reproduction number and infections. From Figures 8 (a) and (c), it is apparent that *ℛ*_*m*_ increases when *b*_*m*_ increases and *ℛ*_*m*_ decreases when *µ*_*v*_ increases. Figures 8 (b) and (d) follow the similar trend. For instance, Figure 8 (b) shows that the daily infection level cases increases by around 150 % when *b*_*m*_ increases from 0.2 to 0.4. However, Figure 8 (d) depicts almost a 40 % decrease in the highest daily infection count when *µ*_*v*_ increases from 0.12 to 0.18. The effect of fully recovery from malaria and recovered carriers are also assessed in Figure 9. Figures 9 (a) and (b) illustrate that if all the individuals recover fully from malaria, *ℛ*_*m*_ remains less than one. Hence, the disease will be under control. Further, Figures 9 (c) and (d) demonstrate that when individuals recover from malaria and at the same time carry the parasite, *ℛ*_*m*_ becomes greater than one. Hence, malaria will persist in the community. The model is further analyzed through simulation to evaluate the effect of both the drug-resistant and drug-sensitive strains on malaria dynamics by calculating infection to recovery ratio. According to Figure 10 (a), individuals infected with the drug-resistant strain exhibit a recovery rate of about 22%, whereas Figure 10 (b) indicates a recovery rate of approximately 42% for those infected with the drug-sensitive strain. Now, we will discuss the effect of various optimal control strategies on the spread of malaria.

**Table 1.**
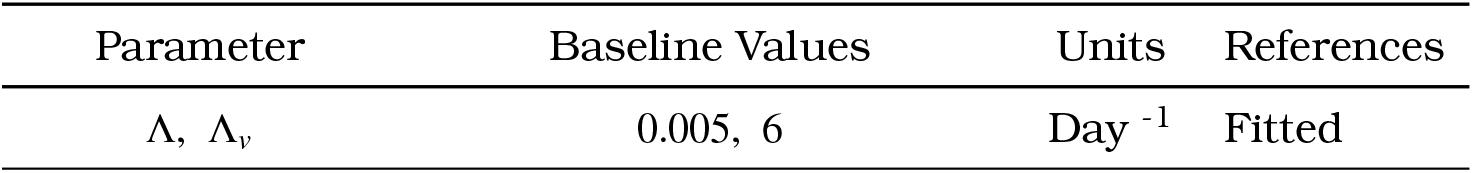
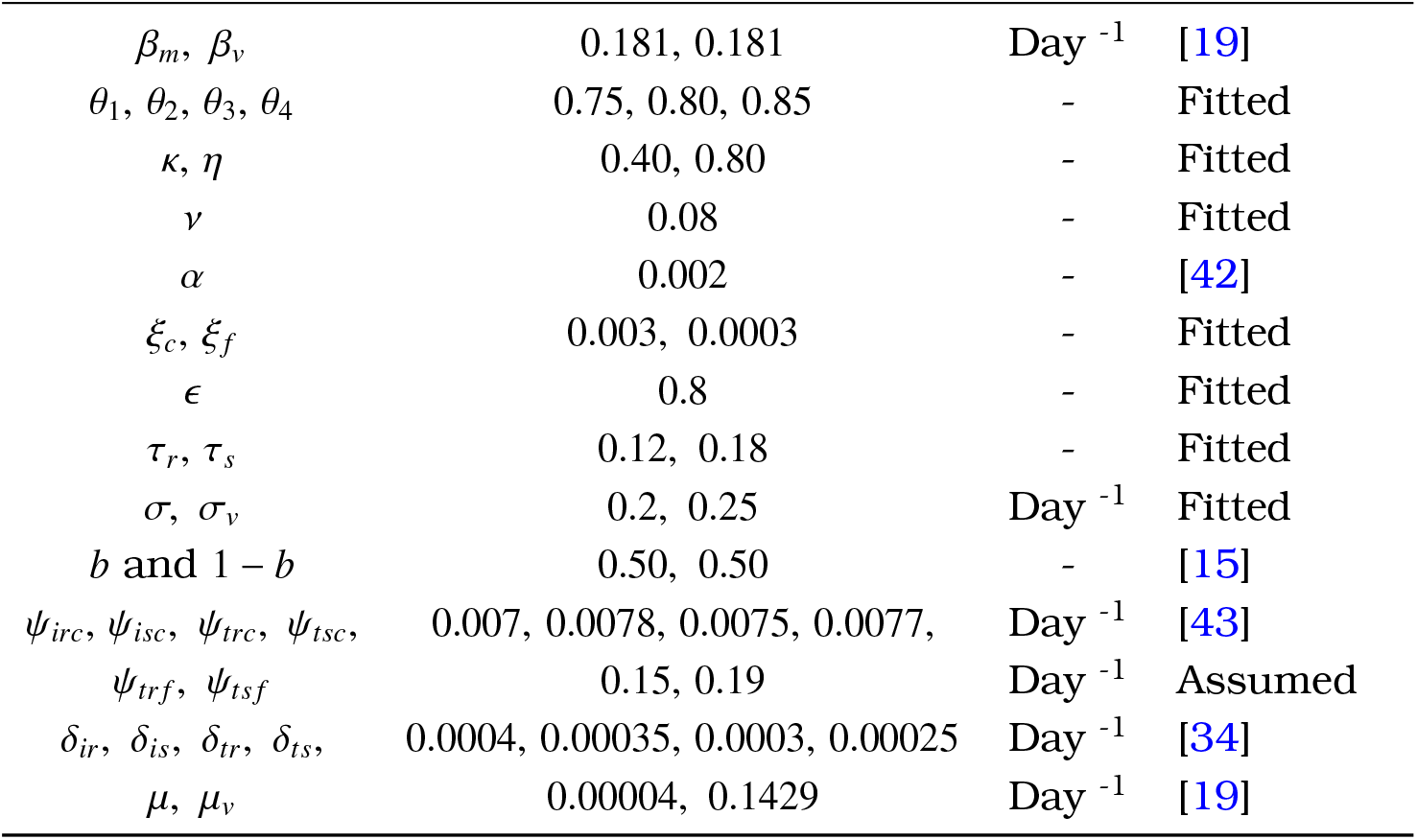
Numerical values of the parameters for the model (13).

**Figure 2.**
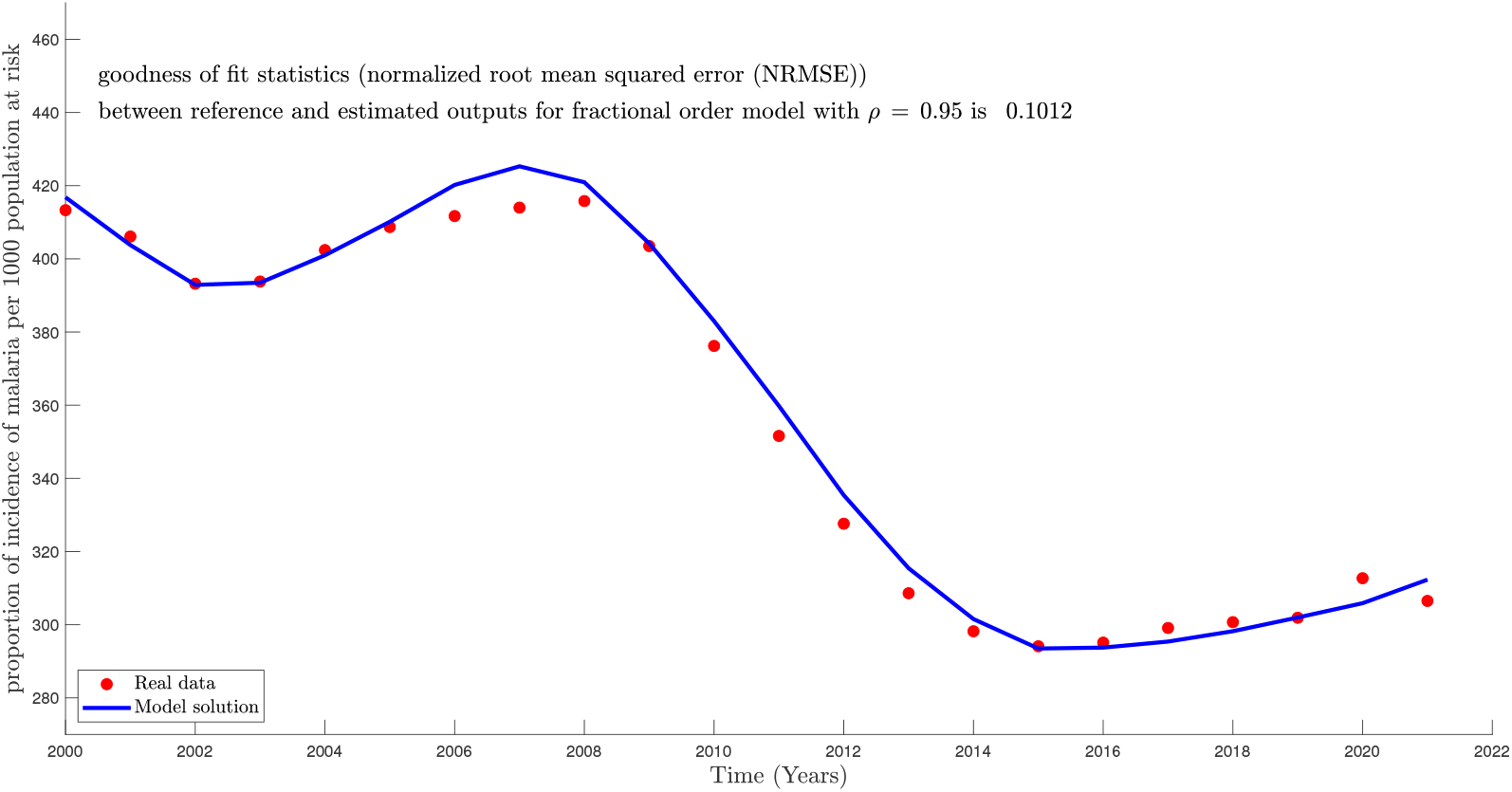
Simulation of the model (13) illustrates the comparison between the real data and model output.

**Figure 3.**
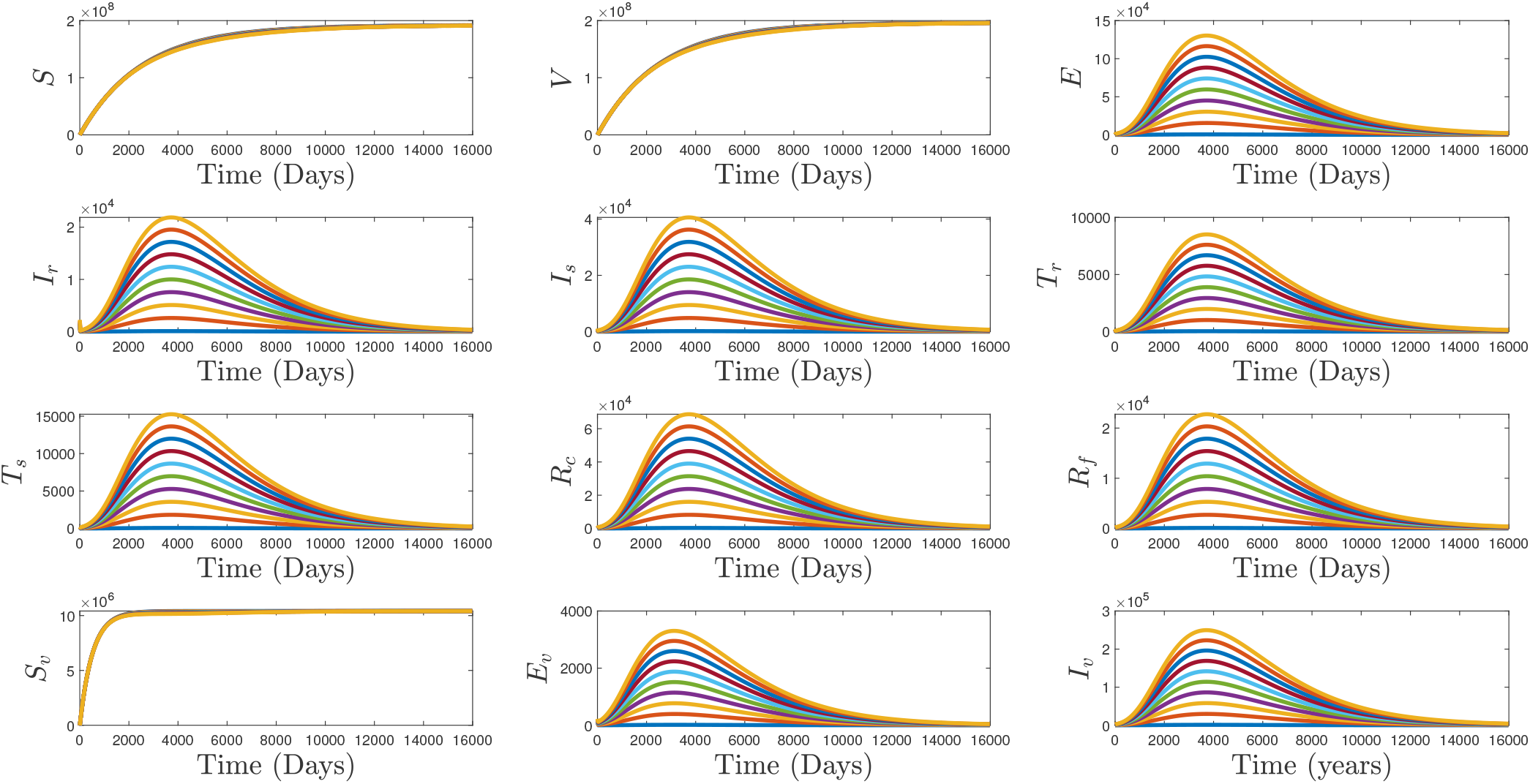
Simulation illustrates that solution trajectories tend to DFE when *ℛ*_*m*_ *<* 1.

**Figure 4.**
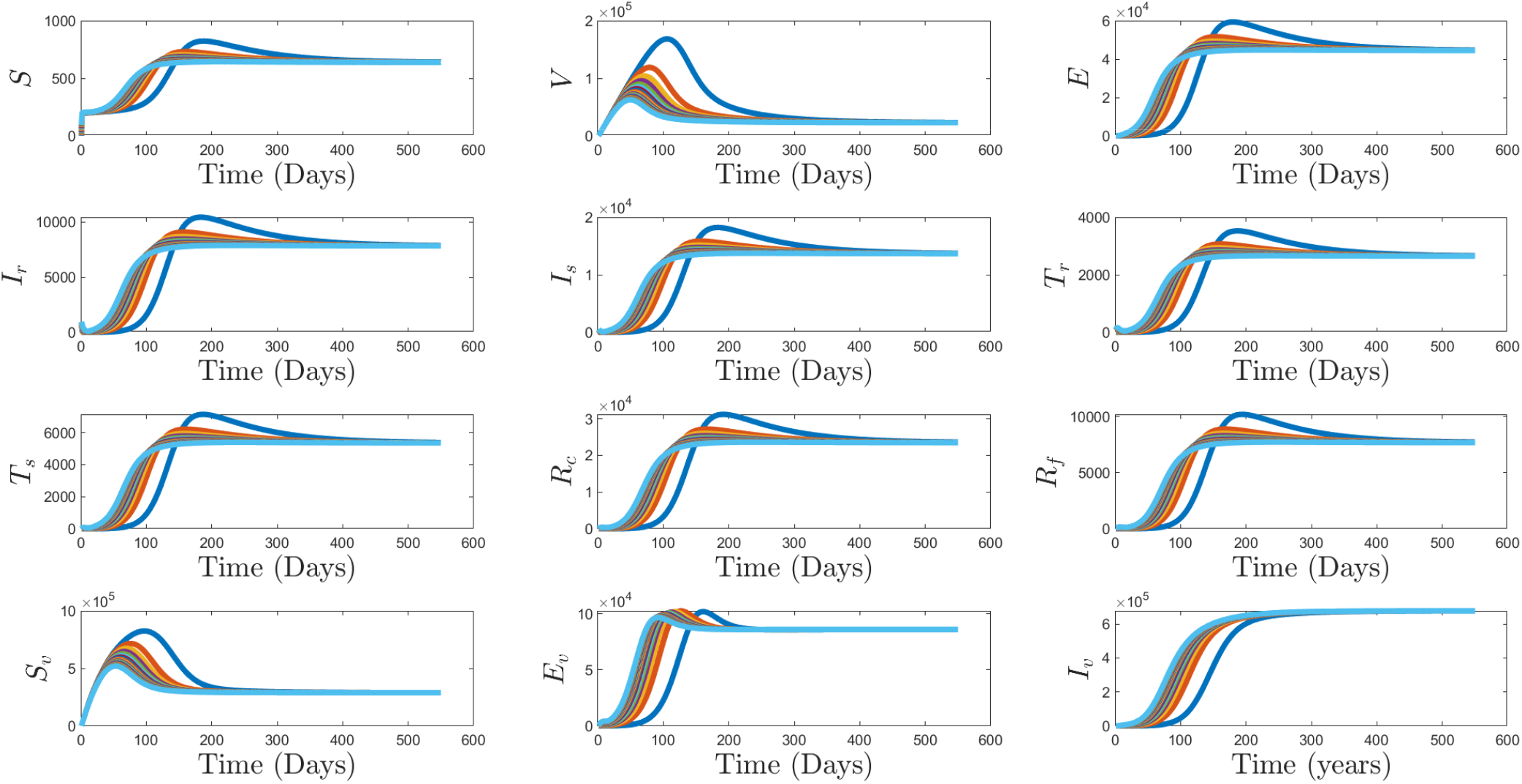
Simulation illustrates that solution trajectories tend to EEP when *ℛ*_*m*_ *>* 1.

**Figure 5.**
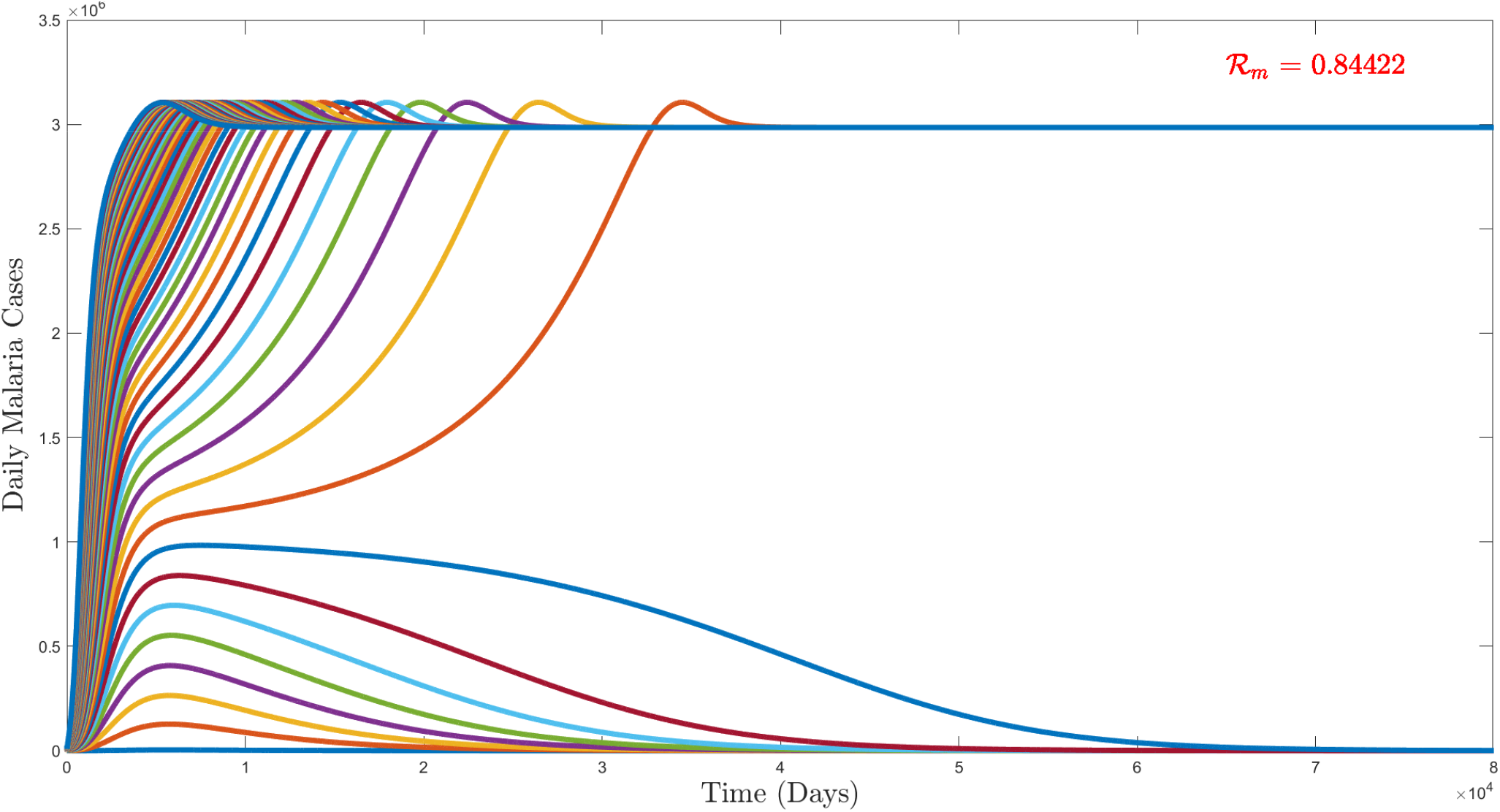
Numerical simulation presents the existence of backward bifurcation as solution trajectories tend to both DFE and EEP when *ℛ*_*m*_ *<* 1.

**Figure 6.**
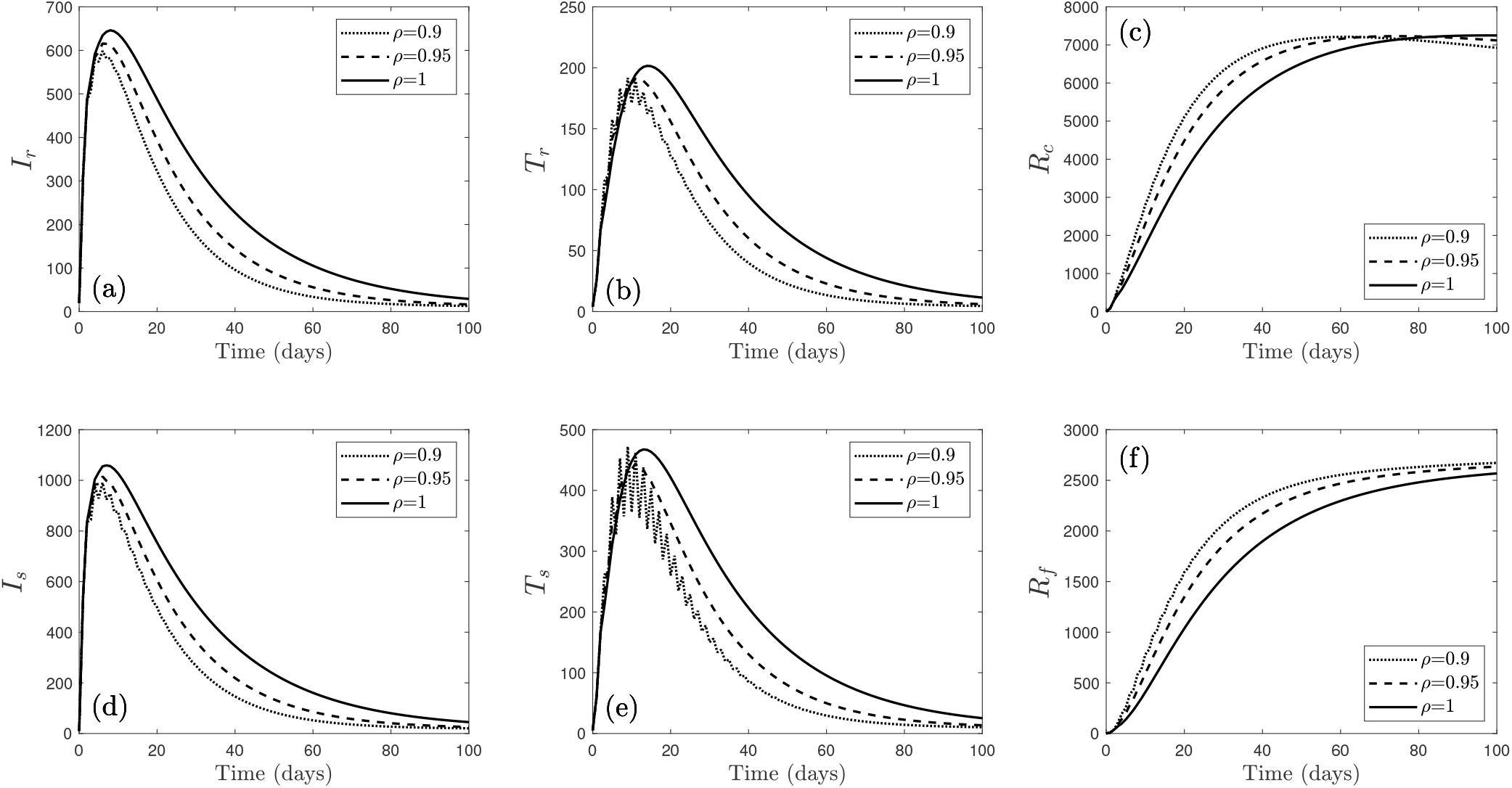
Model output of Infected, Treated and Recovered classes, respectively obtained from simulation of the model (13).

**Figure 7.**
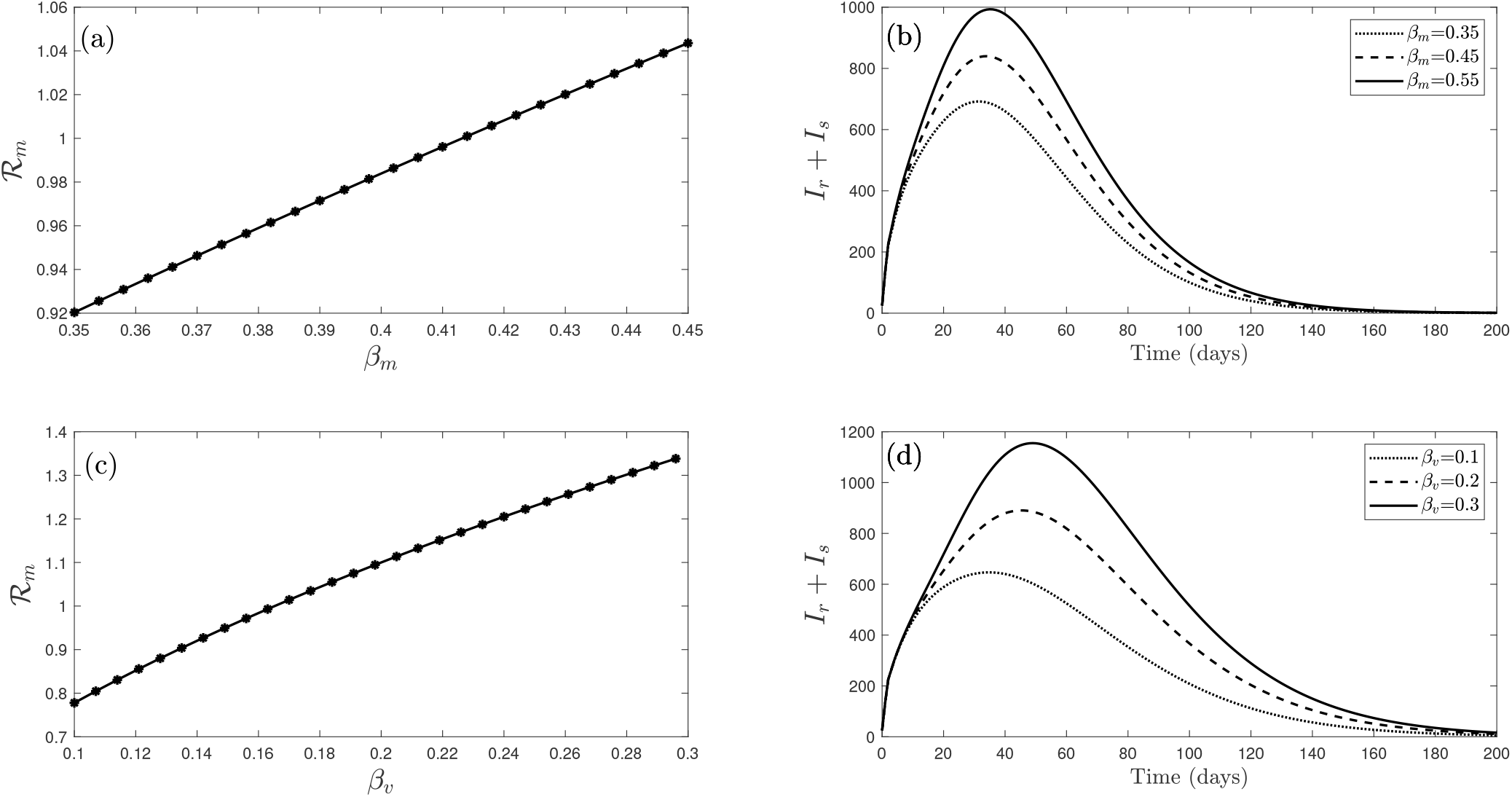
Visual presentation of the model (13) depicts the effect of *β*_*m*_ and *β*_*v*_ on reproduction number and infected cases.

**Figure 8.**
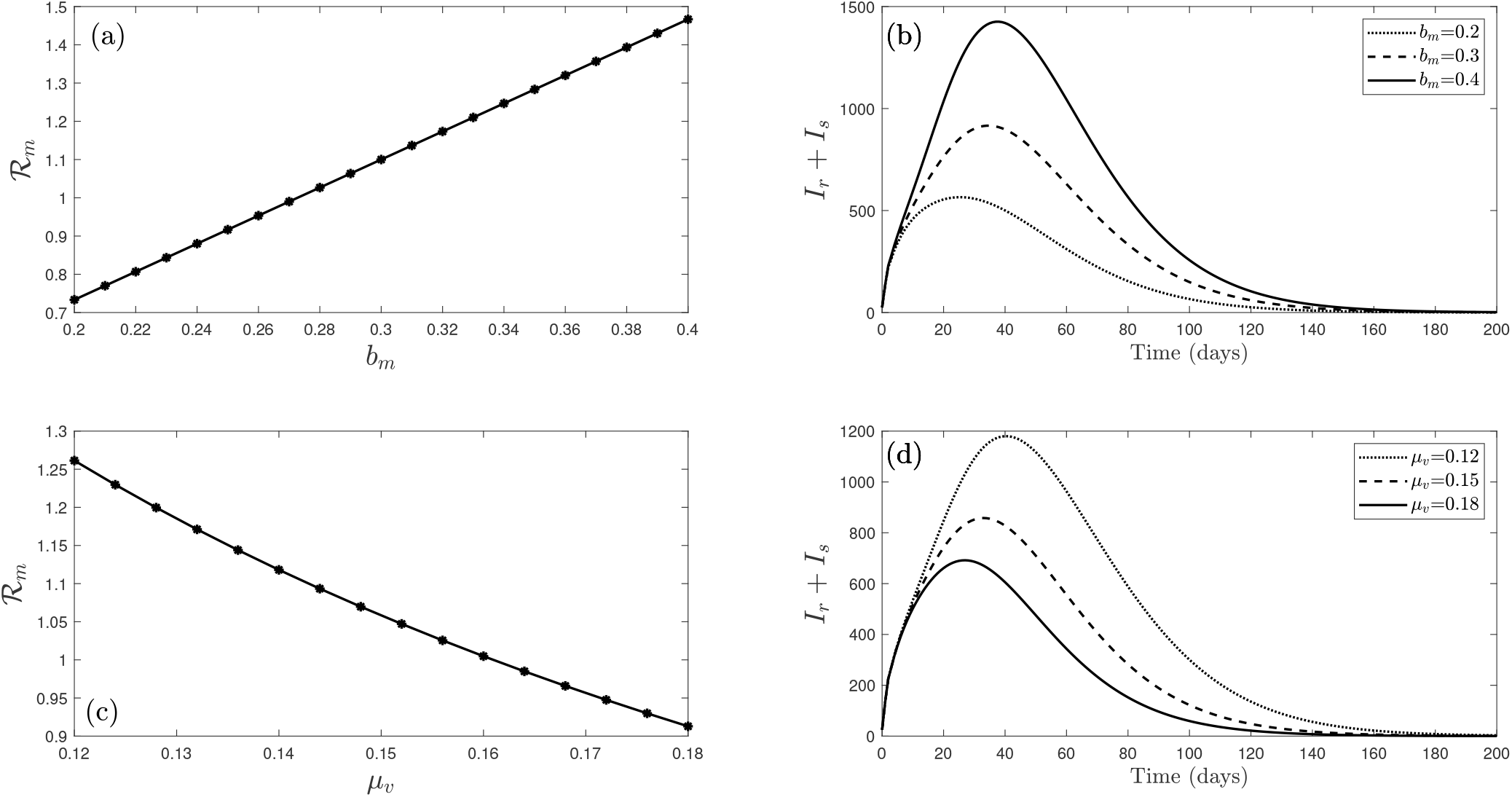
Visual presentation of the model (13) shows the impact of *µ*_*v*_ and *b*_*m*_ on *ℛ*_*m*_ and infected class.

**Figure 9.**
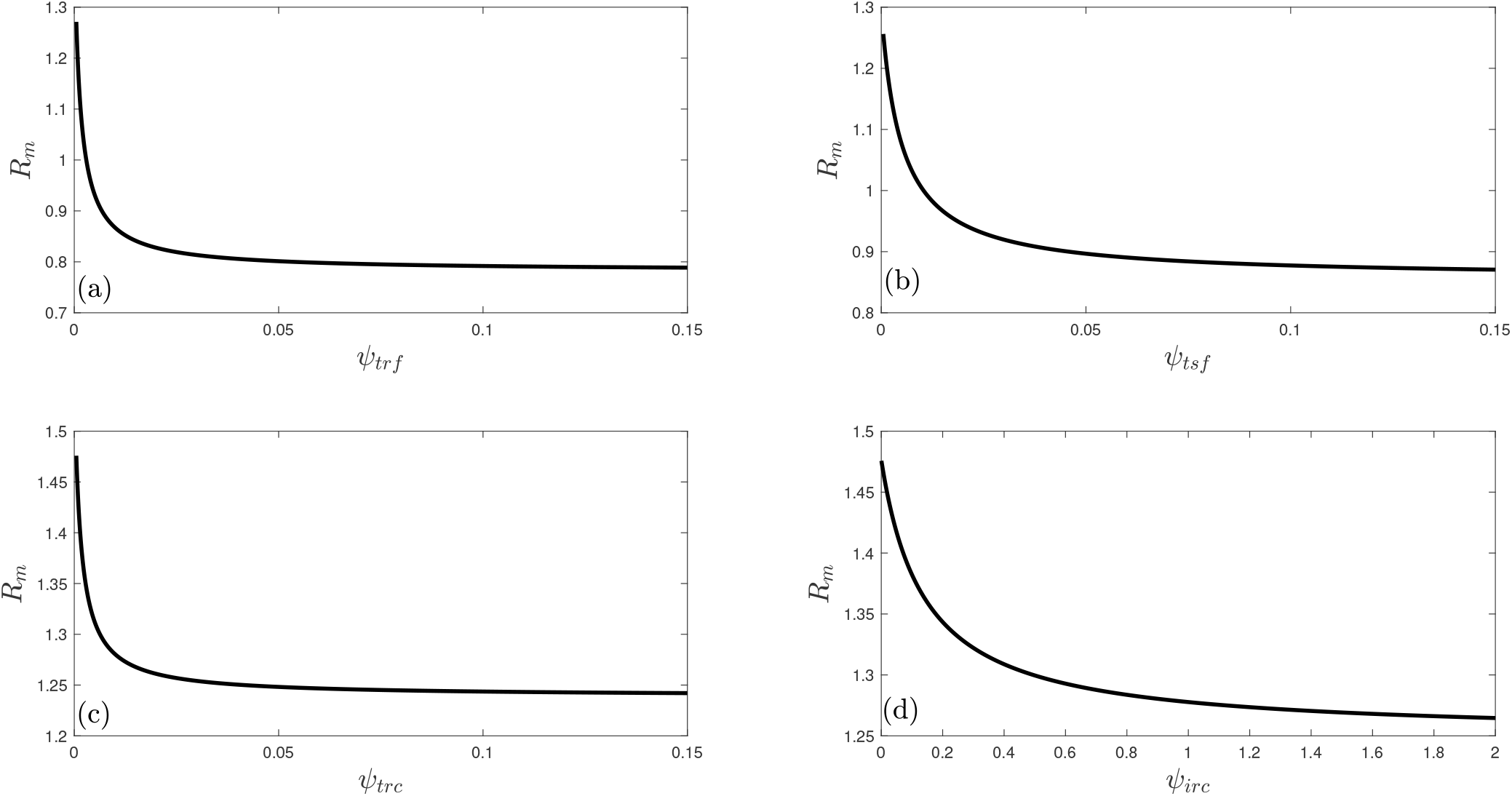
Numerical simulation of the model (13) shows the effect of full recovery from malaria and recovered carrier on *ℛ*_*m*_.

**Figure 10.**
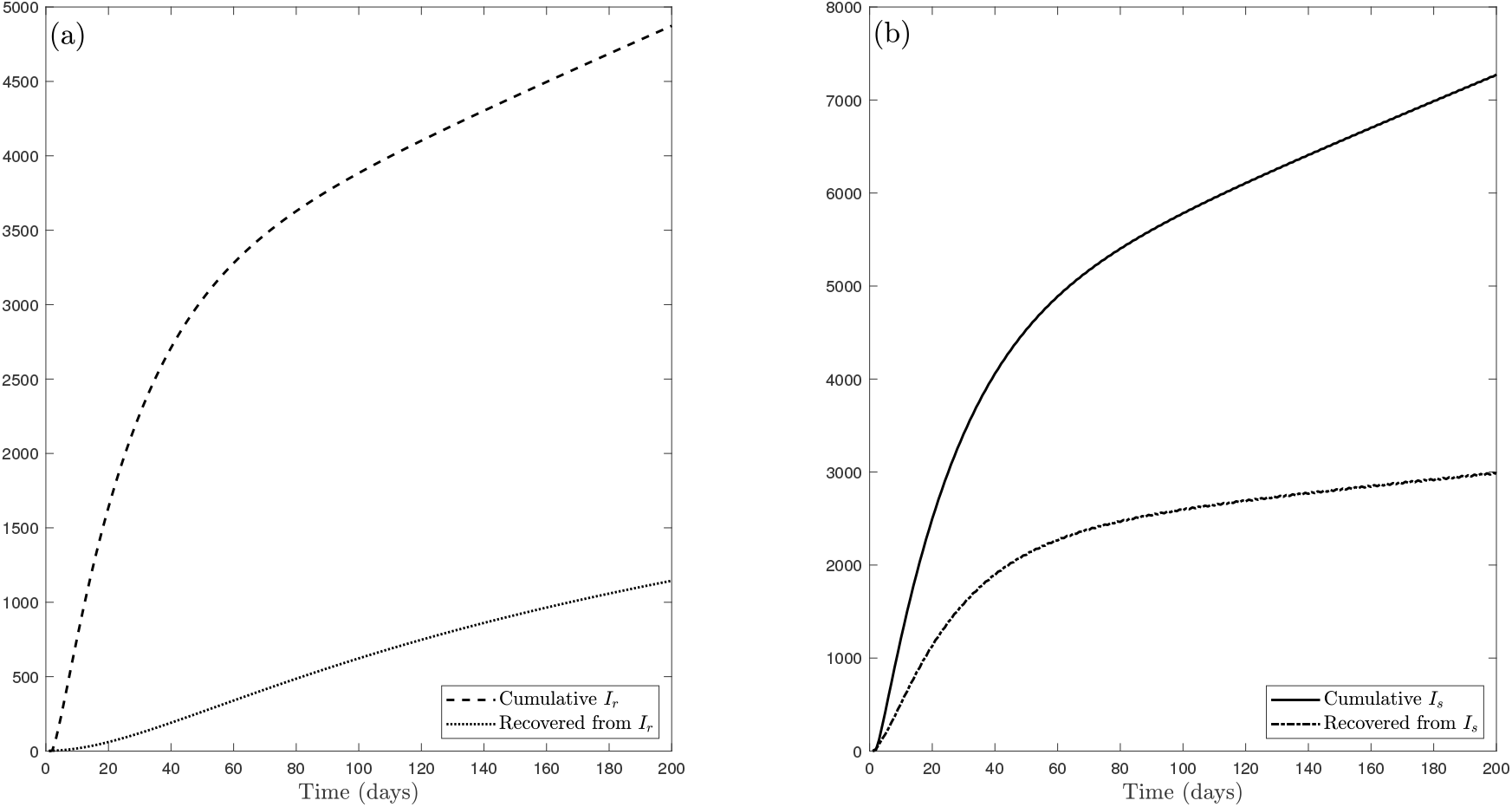
Graphical representation of the model (13) displays the effect of both drug-resistant and drug-sensitive strains on the dynamics of the disease.

### 7.1 Strategy A: Malaria prevention control (u_1_)

In this strategy, malaria prevention control (*u*_1_) is applied while other controls (*u*_2_ and *u*_3_) are set to zero. Figure 11 (b) depicts a significant reduction in malaria-infected cases. For example, control *u*_1_ averts approximately 11, 000 new malaria cases. Since this strategy prevents individuals from being infected with malaria, it increases susceptible population (Figure 11 (a)) and reduces recovered population (Figure 11 (c)) (as less infection implies less recovery). Figure 11 (d) reflects the change of the control profile for the control variable *u*_1_. This figure confirms that control *u*_1_ should be kept at the maximum strength for approximately the first 73 days after which it gradually decreases to zero.

**Figure 11.**
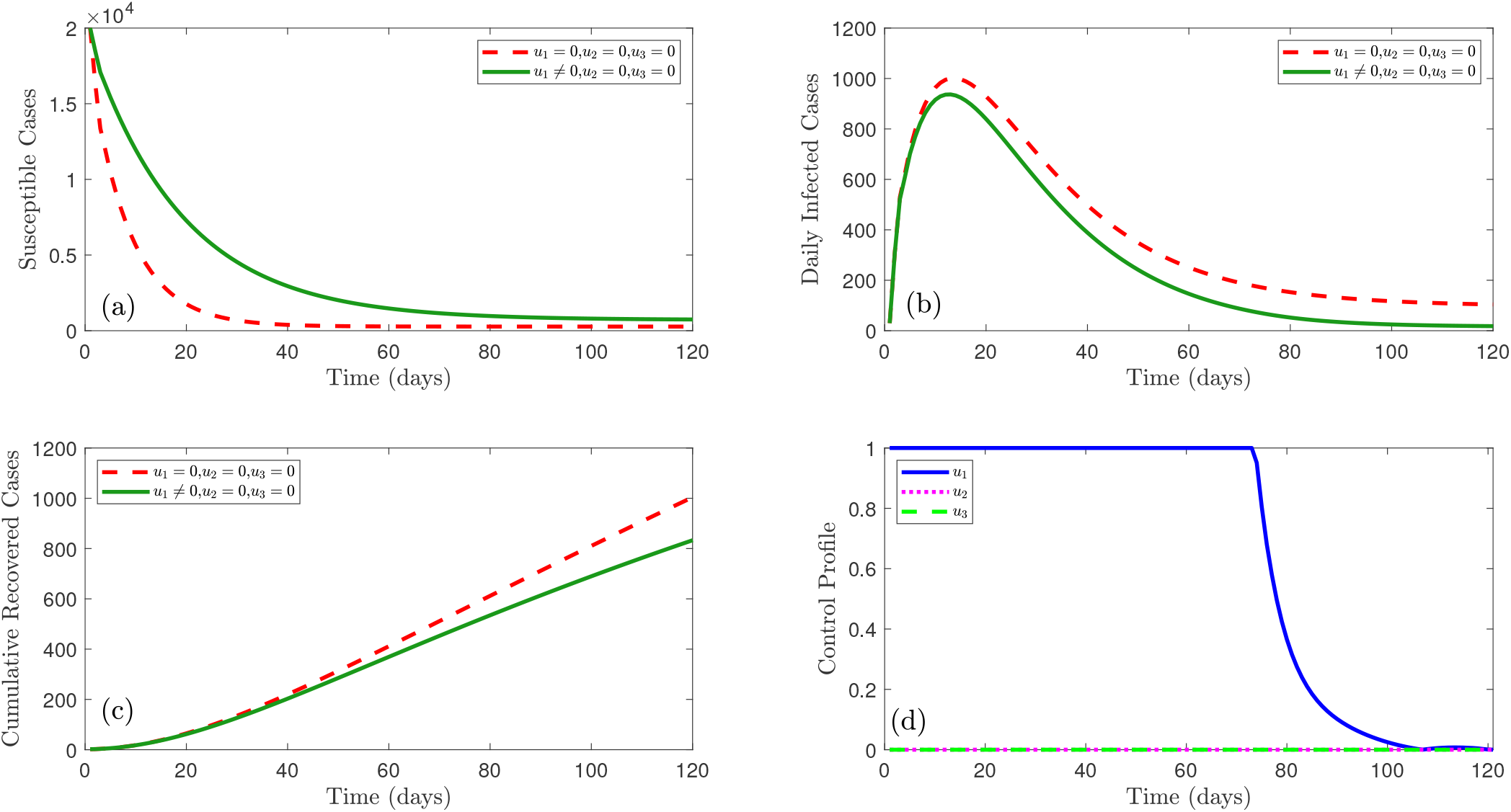
Effect of malaria prevention control (*u*_1_) on the susceptible individuals, infected individuals, and recovered individuals, respectively.

### 7.2 Strategy B: Malaria vaccination control (u_2_)

In this strategy, malaria vaccination control (*u*_2_) is applied to ensure continuous vaccination and increase vaccination campaigns. Other controls (*u*_1_ and *u*_3_) are set to zero. From Figure 12 (b) we observe that this control averts malaria-infected cases significantly. For example, control *u*_2_ prevents almost 8, 700 new malaria cases. Like the control *u*_1_, this control also increases susceptible population (Figure 12 (a)) and reduces recovered individuals (Figure 12 (c)). Figure 12 (d) indicates that control *u*_2_ should remain at its peak intensity for about the initial 93 days before decreasing to zero.

**Figure 12.**
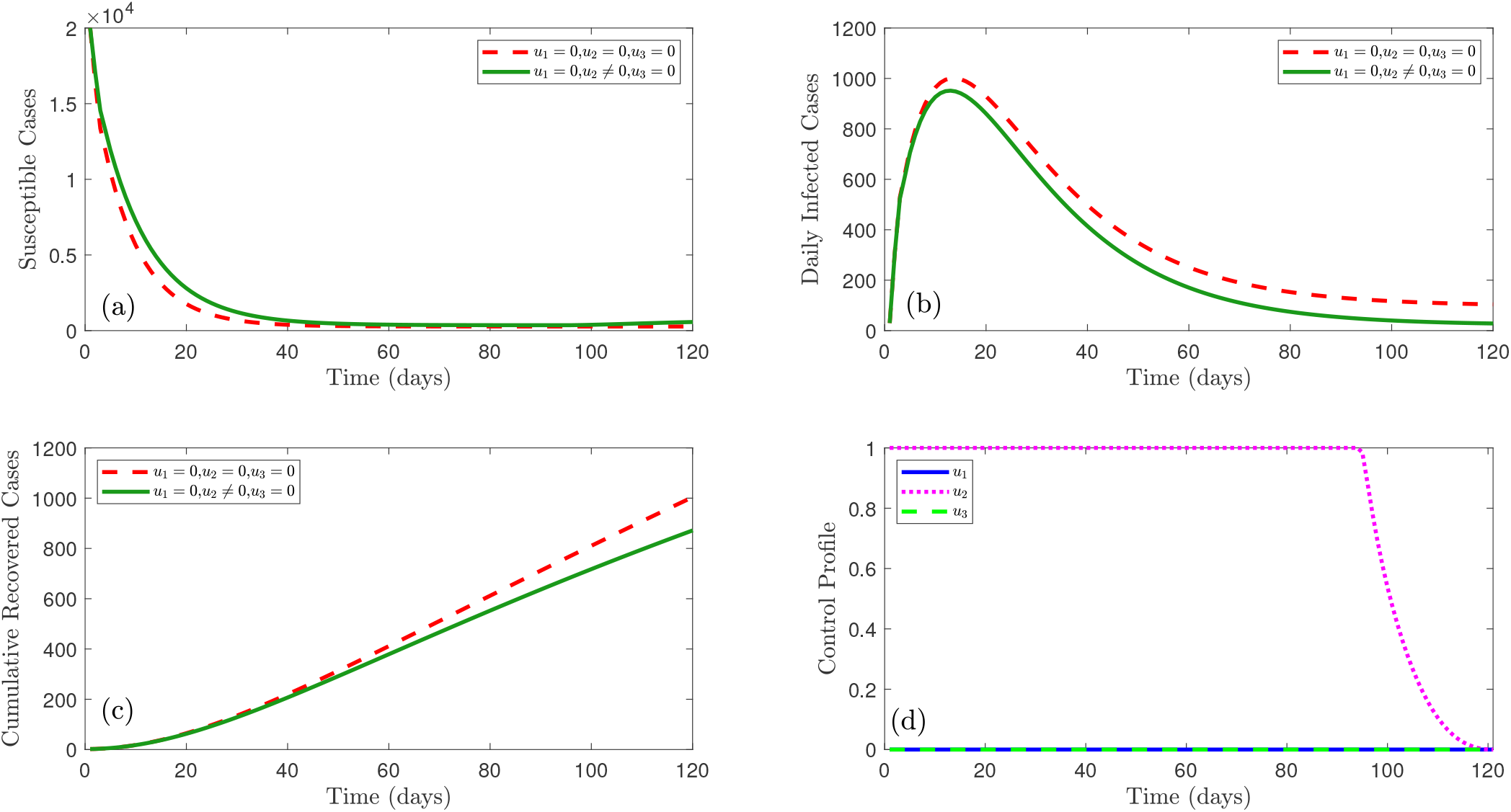
Effect of malaria vaccination control (*u*_2_) on the susceptible individuals, infected individuals, and recovered individuals, respectively.

### 7.3 Strategy - C: Malaria treatment control (u_3_)

In this strategy, malaria treatment control (*u*_3_) is used to ensure better treatment for infected individuals while other controls (*u*_1_ and *u*_2_) are set to zero. Figure 13 (b) depicts that this control removes approximately 8, 200 malaria cases. Furthermore, this control increases the number of susceptible individuals (Figure 13 (a)) and recovered individuals (Figure 13 (c)). From the control profile (Figure 13 (d)) it is observed that control *u*_3_ should be maintained at the peak level for almost all the time period.

**Figure 13.**
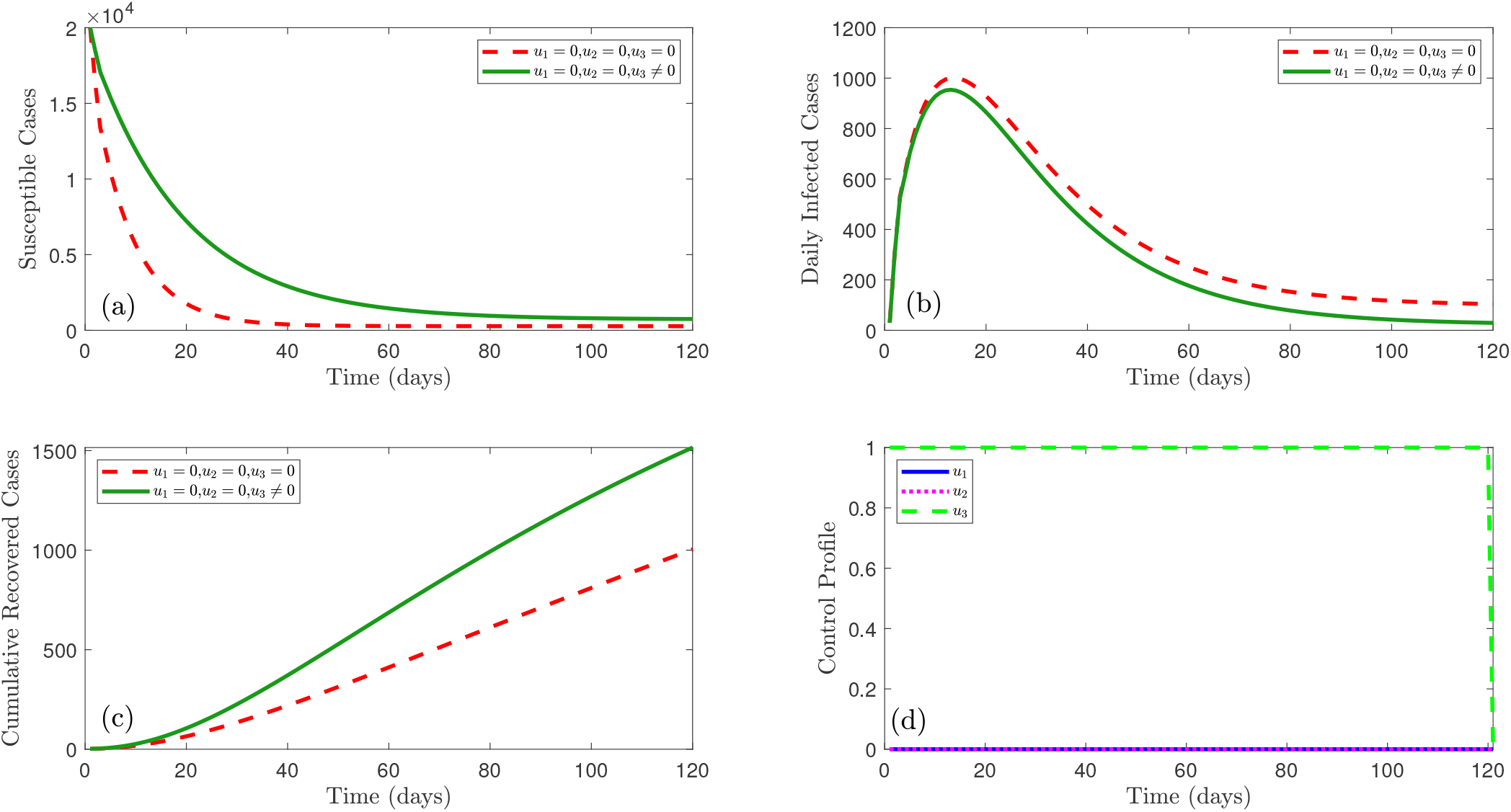
Effect of malaria treatment control (*u*_3_) on the susceptible individuals, infected individuals, and recovered individuals, respectively.

### 7.4 Strategy - D: Combination of malaria prevention control (u_1_) and vaccination control (u_2_)

In this strategy, a combination of malaria prevention control (*u*_1_) and vaccination control (*u*_2_) is used to reduce disease burden. Other control (*u*_3_) is set to zero. Figure 14 (b) shows that this control averts nearly 11, 050 new malaria cases. According to this figure, this control increases susceptible individuals (Figure 14 (a)) and reduces recovered individuals (Figure 14 (c)). From the control profile (Figure 14 (d)) we see that control *u*_1_ should be applied at full strength for around the first 73 days followed by a gradual reduction to zero. This figure also shows that the strength of control *u*_2_ should be increased gradually from 0.2 and then attain the highest intensity on day 73 and should be kept at the maximum intensity for 8 days before decreasing to zero gradually.

**Figure 14.**
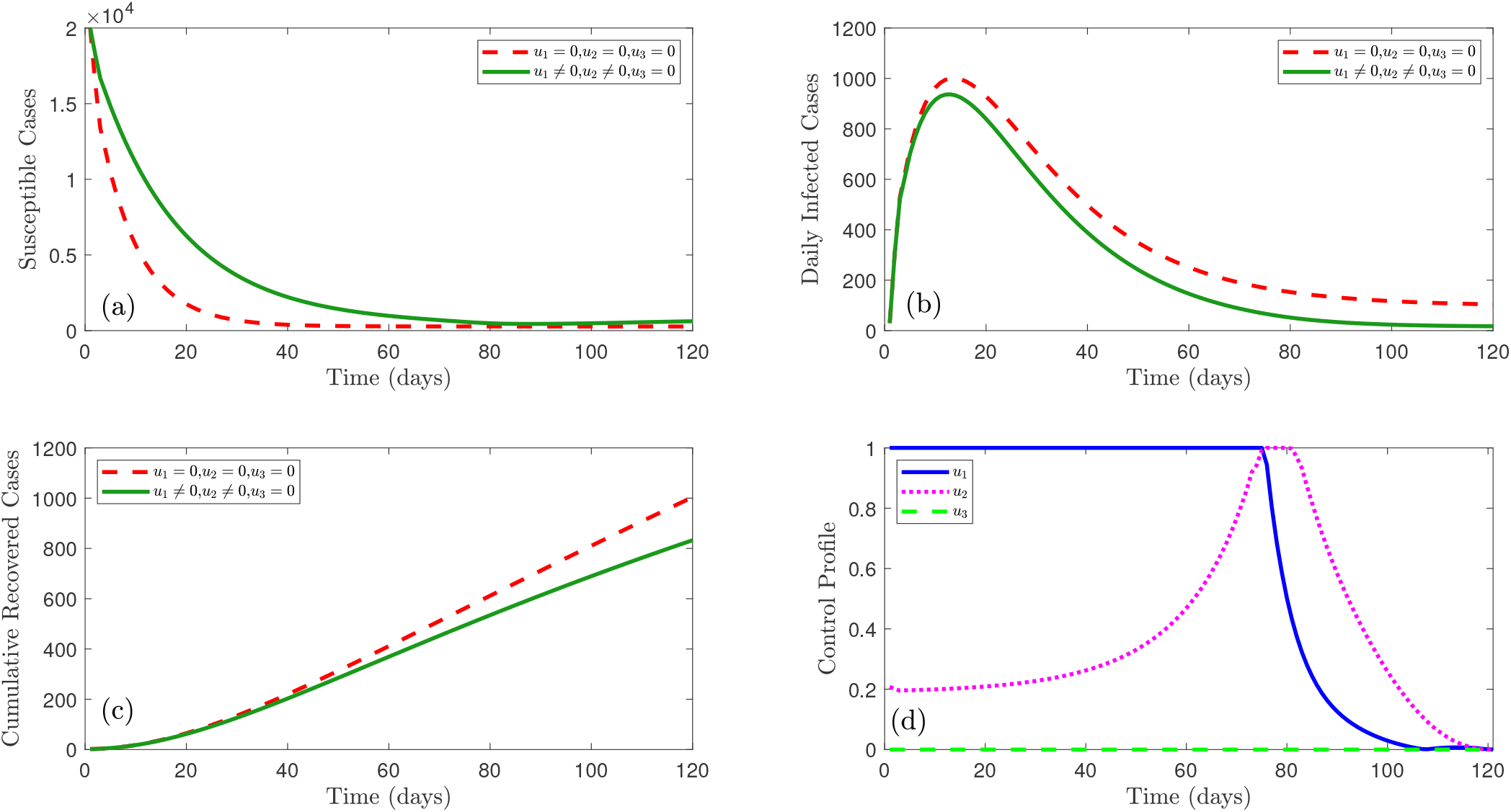
Combined effect of malaria prevention control and vaccination control (*u*_1_ and *u*_2_) on the susceptible individuals, infected individuals, and recovered individuals, respectively.

### 7.5 Strategy - E: Combination of malaria prevention control (u_1_) and treatment control (u_3_)

In this strategy, we use a combination of malaria prevention control (*u*_1_) and treatment control (*u*_3_). Other control (*u*_2_) is set to zero. This strategy reduces the malaria infection (Figure 15 (b)) by almost 11020. This control also increases susceptible individuals (Figure 15 (a)) and recovered individuals (Figure 15 (c)). Control profile (Figure 15 (d)) suggests that control *u*_1_ should remain at the maximum level for about the first 73 days after which it decreases to zero. This figure also shows that the strength of control *u*_3_ should be maintained at the highest intensity for all the time periods.

**Figure 15.**
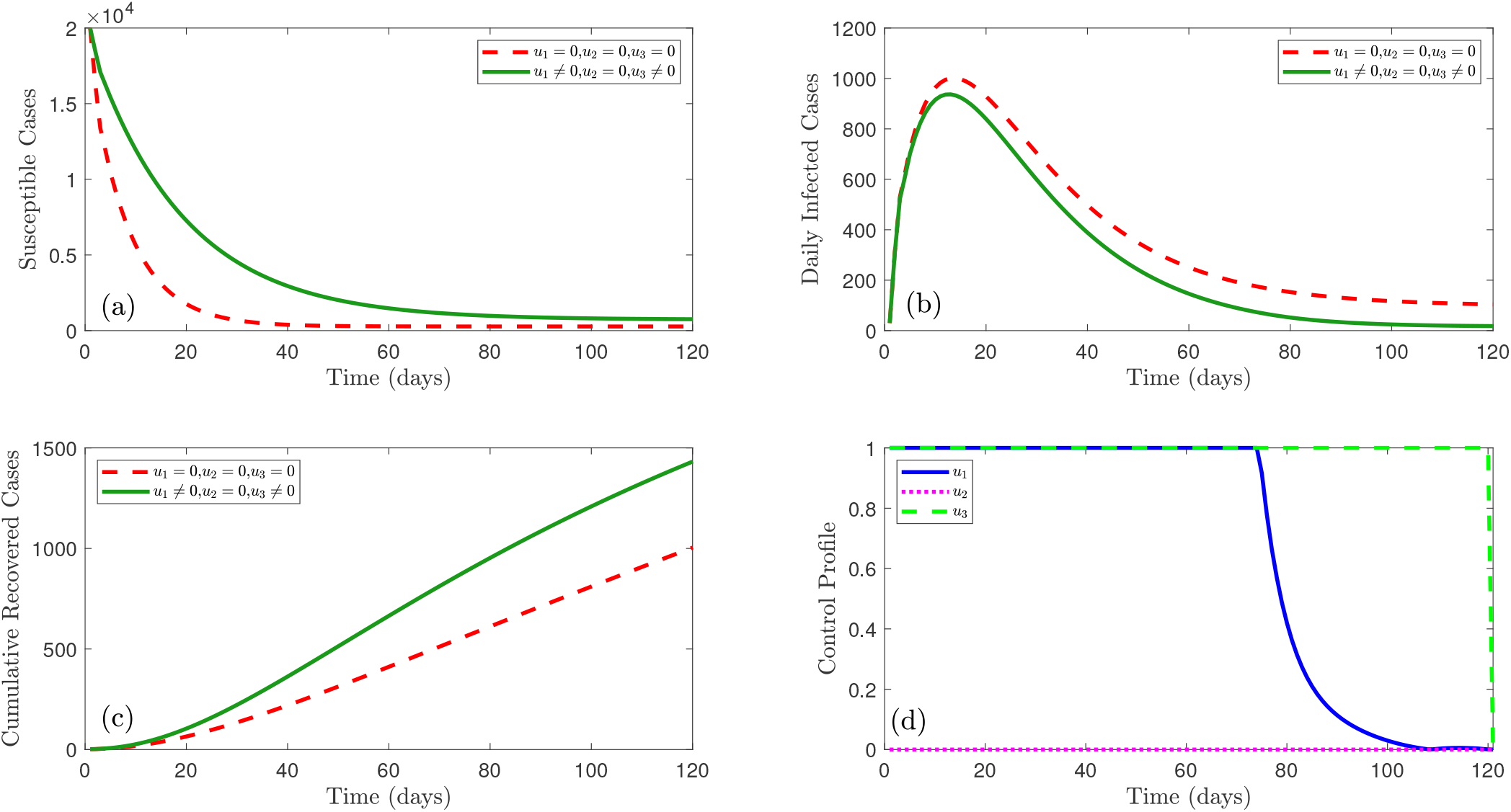
Combined effect of malaria prevention control and treatment control (*u*_1_ and *u*_3_) on the susceptible individuals, infected individuals, and recovered individuals, respectively.

### 7.6 Strategy - F: Combination of malaria vaccination control (u_2_) and treatment control(u_3_)

In this strategy, we apply malaria vaccination control (*u*_2_) and treatment control (*u*_3_) together. Other control (*u*_1_) is set to zero. This strategy reduces the number of infections (Figure 16 (b)) by nearly 8750. It also increases susceptible individuals (Figure 16 (a)) and recovered individuals (Figure 16 (c)). Figure 16 (d) shows that control *u*_2_ stays at the peak strength for a period of approximately 93 days before reducing to zero. This figure also shows that the strength of control *u*_3_ should be maintained at the highest intensity for all the time periods.

**Figure 16.**
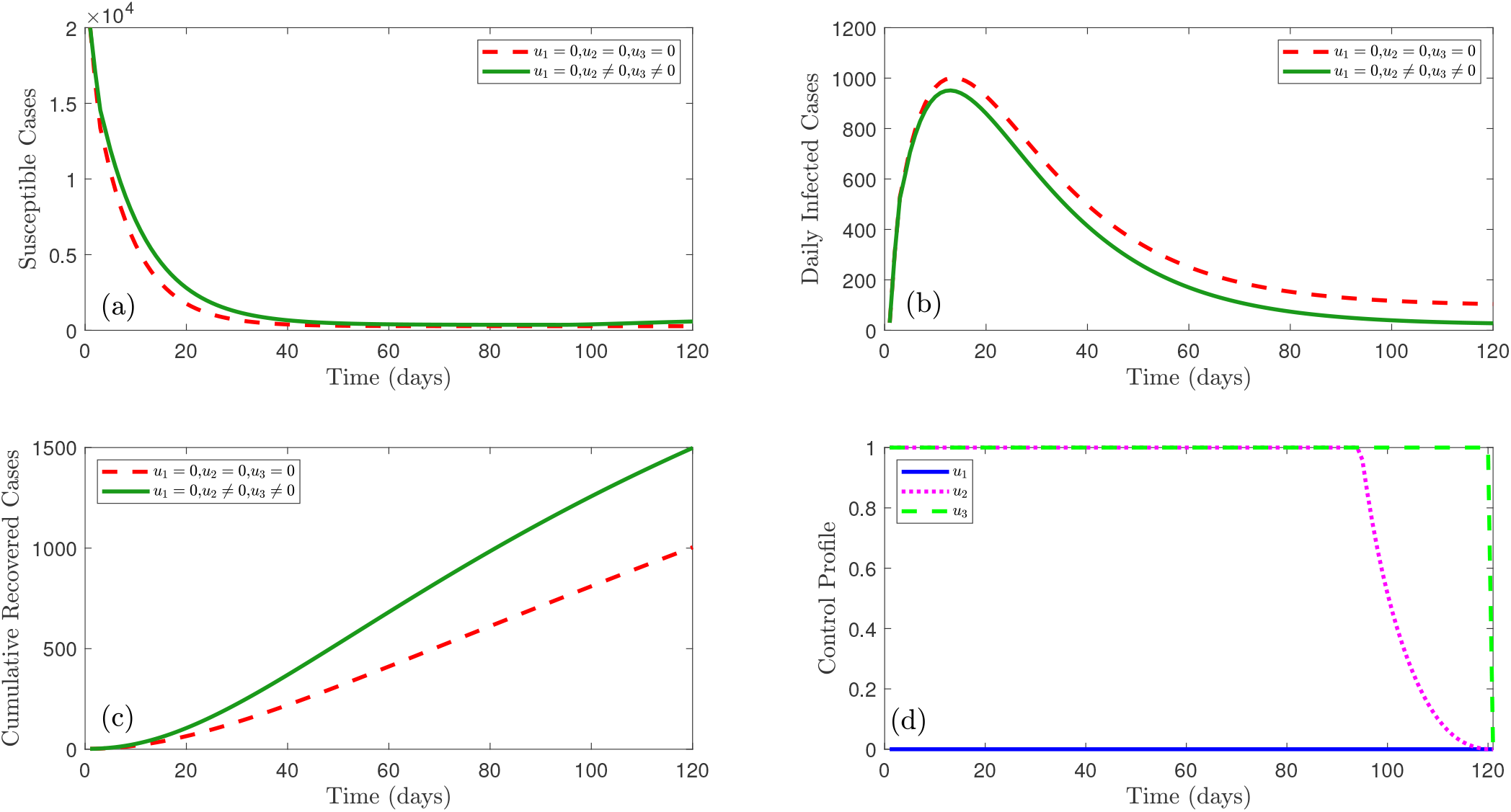
Combined effect of malaria vaccination control and treatment control (*u*_2_ and *u*_3_) on the susceptible individuals, infected individuals, and recovered individuals, respectively.

### 7.7 Strategy - G: Combination of malaria prevention control (u_1_), vaccination control (u_2_) and treatment control (u_3_)

In this strategy, we have considered all the controls. The difference in the number of susceptible cases, infected cases, and recovered cases between no control and this strategy is shown in Figure 17. Using this strategy the number of infected cases (Figure 17 (b)) can be reduced by about 24 %. Number of susceptible individuals (Figure 17 (a)) and recovered individuals (Figure 17 (c)) increase by approximately 105 % and 53 %, respectively. Figure 17 (d) suggests that control *u*_1_ should be kept at the maximum for nearly 73 days and then gradually reduces to zero. This figure also shows that control *u*_2_ increases gradually from 0.2 and by around 73 days reaches the maximum intensity, holds this level for almost 8 days and then gradually decreases to zero. Further, it is observed that control *u*_3_ should be maintained at the highest intensity for all the time periods.

**Figure 17.**
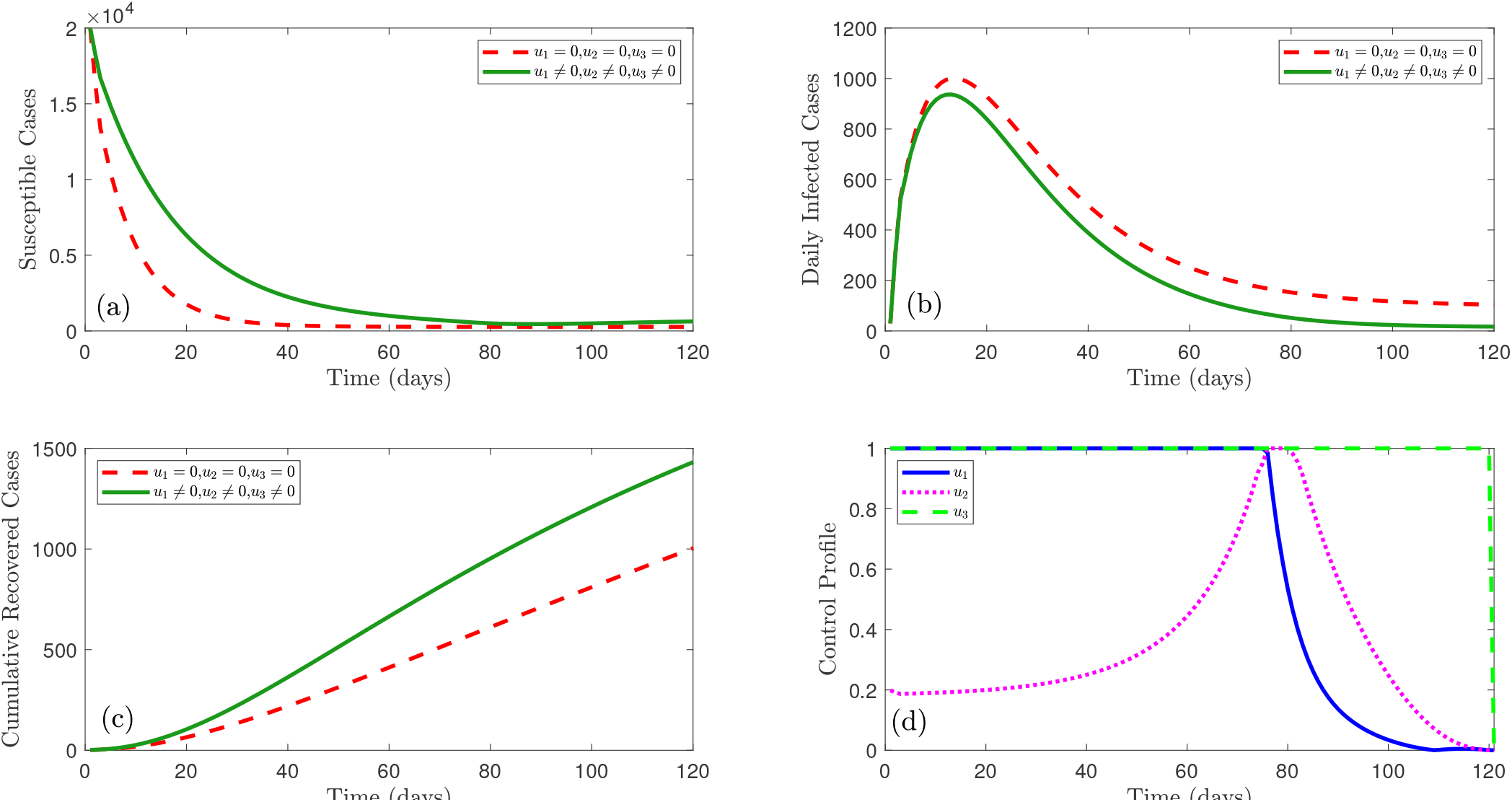
Combined effect of all controls (*u*_1_, *u*_2_, and *u*_3_) on the susceptible individuals, infected individuals, and recovered individuals, respectively.

### 7.8 Cost-effectiveness analysis

In section 6, we have analyzed the optimal control model considering three control measures, namely, prevention control (*u*_1_), vaccination control (*u*_2_), and treatment control (*u*_3_). Further, in section 7, graphical representation regarding the optimal control analysis is provided. From the numerical simulation, we observe that strategy - G averts the most infected cases, which is followed by strategy - D, strategy - E, strategy - A, strategy - F, strategy - B, strategy - C. However, this data does not indicate which strategy is the most effective, and this necessitates the study of a cost-effectiveness analysis [45, 46]. Seven different control strategies have been applied with cost weights assuming *A*_1_ = 40, *A*_2_ = 0.5, *A*_3_ = 45. Now, we define the average cost-effectiveness ratio (ACER) and the incremental cost-effectiveness ratio (ICER) as in [35].

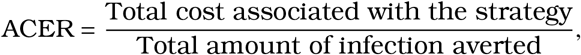

and

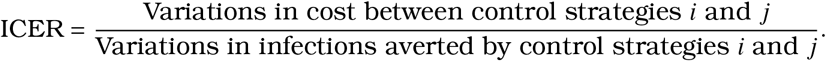

The numerator of ICER represents the difference in cost values, for example, prevention costs, treatment costs, and vaccination costs. The denominator indicates changes in health outcomes, such as infections aversion and the change in recovered individuals. The seven control strategies described above are arranged in ascending order based on the number of infections averted. After that, usning the above definition, ACER and ICER are calculated for each strategy and shown in table 2(a). From this table it is observed that strategy - G has the largest ICER value and hence is the most expensive and least effective strategy in reducing disease complexity. Hence this strategy is omitted from the list and the updated list is provided in table 2(b). ACER and ICER are then calculated for table 2(b). The table shows that strategy - E should be removed from the list as it has the largest ICER value which makes it less effective. The new list has been shown in table 3(a). Following the same procedure, from table 4(b) it can be concluded that Strategy - B is the most cost-effective strategy to eliminate malaria from the community.

**Table 2.** Table 2(a) and table 2(b) presents the results for the 1^st^ and 2^nd^ steps corresponding to (53).

| Strategy | Infection Averted | Total Cost | ACER | ICER |
| --- | --- | --- | --- | --- |
| C | 8188 | 45 | 0.00550 | 0.00550 |
| B | 8694 | 0.5 | 0.00006 | -0.08794 |
| F | 8752 | 45.5 | 0.00520 | 0.77586 |
| A | 11008 | 40 | 0.00363 | -0.00244 |
| E | 11020 | 85 | 0.00771 | 3.75 |
| D | 11047 | 40.5 | 0.00367 | -1.64815 |
| G | 11052 | 85.5 | 0.00774 | 9 |

| Strategy | Infection Averted | Total Cost | ACER | ICER |
| --- | --- | --- | --- | --- |
| C | 8188 | 45 | 0.00550 | 0.00550 |
| B | 8694 | 0.5 | 0.00006 | -0.08794 |
| F | 8752 | 45.5 | 0.00520 | 0.77586 |
| A | 11008 | 40 | 0.00363 | -0.00244 |
| E | 11020 | 85 | 0.00771 | 3.75 |
| D | 11047 | 40.5 | 0.00367 | -1.64815 |

**Table 3.** Table 3(a) and table 3(b) presents the results for the 3^rd^ and 4^th^ steps corresponding to (53).

| Strategy | Infection Averted | Total Cost | ACER | ICER |
| --- | --- | --- | --- | --- |
| C | 8188 | 45 | 0.00550 | 0.00550 |
| B | 8694 | 0.5 | 0.00006 | -0.08794 |
| F | 8752 | 45.5 | 0.00520 | 0.77586 |
| A | 11008 | 40 | 0.00363 | -0.00244 |
| D | 11047 | 40.5 | 0.00367 | 0.01282 |

**3(b)** Strategies C, B, A, and D are arranged in ascending order in terms of infection averted
| Strategy | Infection Averted | Total Cost | ACER | ICER |
| --- | --- | --- | --- | --- |
| C | 8188 | 45 | 0.00550 | 0.00550 |
| B | 8694 | 0.5 | 0.00006 | -0.08794 |
| A | 11008 | 40 | 0.00363 | 0.01707 |
| D | 11047 | 40.5 | 0.00367 | 0.01282 |

**Table 4.** Table 4(a) and table 4(b) presents the results for the 5^th^ and 6^th^ steps corresponding to (53).

| Strategy | Infection Averted | Total Cost | ACER | ICER |
| --- | --- | --- | --- | --- |
| C | 8188 | 45 | 0.00550 | 0.00550 |
| B | 8694 | 0.5 | 0.00006 | -0.08794 |
| D | 11047 | 40.5 | 0.00367 | 0.01700 |

**4(b)** Strategies C and B are arranged in ascending order in terms of infection averted
| Strategy | Infection Averted | Total Cost | ACER | ICER |
| --- | --- | --- | --- | --- |
| C | 8188 | 45 | 0.00550 | 0.00550 |
| B | 8694 | 0.5 | 0.00006 | -0.08794 |

## 8. Conclusion

In this paper, we have formulated a novel CFFOD malaria model to better understand the malaria transmission dynamics as it considers the memory effect. Three control variables are considered to assess the effect of control variables in reducing the spread of malaria. Necessary optimality conditions are then derived. Three-step Adams–Bashforth scheme is considered to analyze the model numerically and to provide graphical representation. In numerical simulation, we have examined the validation of our model with the real data of Nigeria obtained from the World Bank from 2000 − 2021 and it is observed that our model fits well with the obtained data. Then the effect of the order of the fraction on the dynamics of malaria is discussed. The result shows that the larger the order the slower the process and vice versa. Numerical simulation and corresponding graphical outcome show that an increase in effective transmission rate and biting rate can enhance number malaria infection. This study also shows that malaria transmission can be brought into under control by killing more mosquitoes. The effect of the presence of drug resistance strain is also shown using numerical simulation. This study reveals that due to the presence of drug-resistance strain, infection to recovery ratio decreases significantly. Numerical computation also shows that if malaria-infected patients can not be fully cured of malaria, the basic reproduction number remains greater than one. It implies that the disease will persist in the community. Finally, optimal control and cost-effectiveness analysis have been performed to identify which control strategies can be useful in controlling malaria outbreak. Our analysis suggests that incorporating highly effective vaccines continuously malaria infection can be reduced significantly and this is also the most cost-effective strategy.

## Data Availability

All data are available from the World Bank database (https://data.worldbank.org/indicator/SH.MLR.INCD.P3)

